# Gene-level analysis of rare variants in 363,977 whole exome sequences identifies an association of *GIGYF1* loss of function with type 2 diabetes

**DOI:** 10.1101/2021.01.19.21250105

**Authors:** Aimee M. Deaton, Margaret M. Parker, Lucas D. Ward, Alexander O. Flynn-Carroll, Lucas BonDurant, Gregory Hinkle, Parsa Akbari, Luca A. Lotta, Regeneron Genetics Center, DiscovEHR Collaboration, Aris Baras, Paul Nioi

## Abstract

Sequencing of large cohorts offers an unprecedented opportunity to identify rare genetic variants and to find novel contributors to human disease. We used gene-based collapsing tests to identify genes associated with glucose, HbA1c and type 2 diabetes (T2D) diagnosis in 363,977 exome-sequenced participants in the UK Biobank. We identified associations for variants in *GCK, HNF1A* and *PDX1*, which are known to be involved in Mendelian forms of diabetes. Notably, we uncovered novel associations for *GIGYF1*, a gene not previously implicated by human genetics, in diabetes. *GIGYF1* predicted loss of function (pLOF) variants associated with increased levels of glucose (0.77 mmol/L increase, p = 4.42 × 10^-12^) and HbA1c (4.33 mmol/mol, p = 1.28 × 10^-14^) as well as T2D diagnosis (OR = 4.15, p= 6.14 ×10^-11^). Multiple rare variants contributed to these associations, including singleton variants. *GIGYF1* pLOF also associated with decreased cholesterol levels as well as an increased risk of hypothyroidism. The association of *GIGYF1* pLOF with T2D diagnosis replicated in an independent cohort from the Geisinger Health System. In addition, a common variant association for glucose and T2D was identified at the *GIGYF1* locus. Our results highlight the role of GIGYF1 in regulating insulin signaling and protecting from diabetes.

**Author Summary:** Genetic studies focused on high impact variants in protein-coding regions of the genome can provide valuable insight into the biology of human disease. As these variants tend to be rare, studying them requires large cohort sizes and methods to aggregate variants that are likely to have a similar biological impact. We studied how rare genetic variants contribute to type 2 diabetes (T2D) using sequencing data from 363,977 participants in the UK Biobank, employing methods to aggregate variants at the level of individual genes. As well as identifying genes known to be involved in inherited forms of diabetes, we uncovered a novel association for *GIGYF1. GIGYF1* loss of function associated with increased risk of T2D and increased levels of the diabetes biomarkers glucose and HbA1c. This association was also seen in an independent dataset. *GIGYF1* encodes a protein that binds a negative regulator of the insulin receptor that has not been well-characterized in the literature. By highlighting the importance of GIGYF1 in modulating insulin signaling these results may lead to new therapeutic approaches for diabetes as well as a new appreciation for *GIGYF1* loss of function as a genetic risk factor for T2D.

## Introduction

Human genetics provides powerful methods for understanding the roles of genes and proteins in disease and can lead to new therapeutic hypotheses and drug targets. Genetic evidence based on sequence variants within coding regions of the genome is better at predicting the efficacy and safety of novel therapeutics than evidence from genome-wide association studies (GWAS), which tend to involve common noncoding variants [1-3]. Among coding variants, predicted loss of function (pLOF) variants are particularly informative in association studies because they establish a direct causal link between reduction in gene function and biological outcomes. Additionally, rare missense variants predicted to be deleterious can provide valuable biological insights [4, 5]. However, interrogation of the effects of such variants is hampered by the rarity of these variants and the cohort sizes needed to identify associations [6]. Exome or whole-genome sequencing of large biobanks coupled with gene-level aggregation of rare high impact variants can help to circumvent these challenges [4]. Biobanks offer a considerable advantage over case-control cohorts as they contain richer phenotyping data which often includes biomarker measurements as well as disease diagnoses. This allows a more complete understanding of the biological consequences of damaging variants in particular genes [7, 8].

Diabetes is a disease that has been extensively studied in traditional array-based GWAS with hundreds of associations identified to date [9-12]. Although these studies have given insight into some of the biological mechanisms contributing to diabetes, most of the reported associations are with variants in non-coding regions, making identification of the causal gene challenging. More recently, exome sequencing has been applied to discover protein-coding variants that alter the risk of developing type 2 diabetes (T2D). Sequencing of 20,791 T2D cases followed by the use of gene-based collapsing tests (to aggregate predicted damaging variants) identified associations of *SLC30A8, MC4R* and *PAM* with T2D diagnosis [5].

Using 363,977 whole exome sequences from the UK Biobank (UKBB) we performed gene-level collapsing tests to examine the association of pLOF and damaging missense variants in ∼17,000 genes with biomarkers of glycemic control, glucose, and glycated hemoglobin (HbA1c), as well as T2D diagnosis.

## Results

### Gene-level associations with glucose, HbA1c and T2D

We used 454,787 whole exome sequences from the UK Biobank (UKBB) to identify rare variants with a minor allele frequency (MAF) ≤1% likely to have functional impact; pLOF variants (i.e. frameshift, stop gain, splice donor or splice acceptor variants) called as high confidence by LOFTEE [13] or missense variants predicted to be damaging (Combined Annotation Dependent Depletion [CADD] score ≥ 25). We identified 726,422 rare pLOF variants affecting 16,477 genes, 58.5% of which were singletons (carried by a single individual), and 2.14 million damaging missense variants in 17,312 genes, 49.6% of which were singletons (Supplementary Table 1).

**Table 1:** Gene-level associations with glucose and HbA1c levels. Association of pLOF or damaging missense variants (CADD score ≥ 25) aggregated per gene with glucose and HbA1c levels. The effect is shown in standard deviations (SD) of transformed values as well as in International Federation of Clinical Chemistry (IFCC) units. CI; confidence interval.

| gene | Variant set | title | pvalue | Effect (SD) | effect (units) | 95% CI - | 95% CI + | units | n carrier measured |
| --- | --- | --- | --- | --- | --- | --- | --- | --- | --- |
| GCK | pLOF | glucose | $1.56 \times 10^{-9}$ | 1.00 | 1.24 | 0.84 | 1.65 | mmol/L | 35 |
| GIGYF1 | pLOF | glucose | $4.42 \times 10^{-12}$ | 0.62 | 0.77 | 0.55 | 0.98 | mmol/L | 121 |
| GCK | missense CADD$\geq 25$ | glucose | $6.15 \times 10^{-11}$ | 0.49 | 0.61 | 0.42 | 0.79 | mmol/L | 173 |
| G6PC2 | missense CADD$\geq 25$ | glucose | $4.62 \times 10^{-83}$ | -0.27 | -0.33 | -0.36 | -0.3 | mmol/L | 5128 |
| GCK | pLOF | HbA1c | $2.64 \times 10^{-17}$ | 1.29 | 8.75 | 6.73 | 10.78 | mmol/mol | 38 |
| GIGYF1 | pLOF | HbA1c | $1.28 \times 10^{-14}$ | 0.64 | 4.33 | 3.23 | 5.43 | mmol/mol | 129 |
| HNF1A | pLOF | HbA1c | $2.14 \times 10^{-7}$ | 0.59 | 4.01 | 2.50 | 5.53 | mmol/mol | 68 |
| TNRC6B | pLOF | HbA1c | $2.36 \times 10^{-7}$ | 0.58 | 3.94 | 2.45 | 5.43 | mmol/mol | 70 |
| RHAG | pLOF | HbA1c | $3.31 \times 10^{-34}$ | -0.86 | -5.81 | -6.75 | -4.88 | mmol/mol | 179 |
| EPB41 | pLOF | HbA1c | $3.14 \times 10^{-17}$ | -0.53 | -3.58 | -4.41 | -2.75 | mmol/mol | 226 |
| PTPRH | pLOF | HbA1c | $4.39 \times 10^{-10}$ | 0.11 | 0.74 | 0.51 | 0.97 | mmol/mol | 2924 |
| APOB | pLOF | HbA1c | $6.94 \times 10^{-8}$ | 0.23 | 1.57 | 1.00 | 2.15 | mmol/mol | 478 |
| PLD1 | pLOF | HbA1c | $2.99 \times 10^{-7}$ | 0.23 | 1.56 | 0.96 | 2.16 | mmol/mol | 438 |
| EPB42 | pLOF | HbA1c | $6.11 \times 10^{-7}$ | -0.31 | -2.08 | -2.90 | -1.26 | mmol/mol | 234 |
| GCK | missense CADD $\geq 25$ | HbA1c | $1.86 \times 10^{-17}$ | 0.56 | 3.83 | 2.94 | 4.71 | mmol/mol | 201 |
| G6PC2 | missense CADD $\geq 25$ | HbA1c | $6.71 \times 10^{-45}$ | -0.18 | -1.21 | -1.38 | -1.04 | mmol/mol | 5574 |
| PFAS | missense CADD $\geq 25$ | HbA1c | $2.09 \times 10^{-8}$ | -0.05 | -0.32 | -0.44 | -0.21 | mmol/mol | 12621 |
| PDX1 | missense CADD $\geq 25$ | HbA1c | $2.54 \times 10^{-7}$ | 0.06 | 0.41 | 0.25 | 0.56 | mmol/mol | 6694 |
| PIEZO1 | missense CADD $\geq 25$ | HbA1c | $1.0 \times 10^{-132}$ | -0.15 | -1.00 | -1.07 | -0.92 | mmol/mol | 26726 |
| AMPD3 | missense CADD $\geq 25$ | HbA1c | $7.28 \times 10^{-34}$ | 0.13 | 0.86 | 0.72 | 1.00 | mmol/mol | 8258 |
| PFKM | missense CADD $\geq 25$ | HbA1c | $2.16 \times 10^{-28}$ | -0.28 | -1.92 | -2.26 | -1.58 | mmol/mol | 1353 |
| ANK1 | missense CADD $\geq 25$ | HbA1c | $3.12 \times 10^{-19}$ | -0.13 | -0.87 | -1.06 | -0.68 | mmol/mol | 4342 |
| PFKL | missense CADD $\geq 25$ | HbA1c | $2.69 \times 10^{-14}$ | 0.10 | 0.68 | 0.50 | 0.85 | mmol/mol | 5245 |
| AXL | missense CADD $\geq 25$ | HbA1c | $4.11 \times 10^{-12}$ | -0.08 | -0.54 | -0.69 | -0.39 | mmol/mol | 6827 |
| LCAT | missense CADD $\geq 25$ | HbA1c | $1.29 \times 10^{-11}$ | -0.27 | -1.82 | -2.34 | -1.29 | mmol/mol | 565 |
| PLA2G12A | missense<br>CADD $\geq 25$ | HbA1c | $5.74 \times 10^{-11}$ | -0.08 | -0.51 | -0.67 | -0.36 | mmol/mol | 6761 |
| PLD1 | missense<br>CADD $\geq 25$ | HbA1c | $1.51 \times 10^{-10}$ | 0.08 | 0.57 | 0.40 | 0.75 | mmol/mol | 5180 |
| APEH | missense<br>CADD $\geq 25$ | HbA1c | $1.86 \times 10^{-10}$ | 0.29 | 1.96 | 1.36 | 2.57 | mmol/mol | 429 |
| TNFRSF13B | missense<br>CADD $\geq 25$ | HbA1c | $4.63 \times 10^{-9}$ | -0.08 | -0.51 | -0.68 | -0.34 | mmol/mol | 5470 |
| BLVRB | missense<br>CADD $\geq 25$ | HbA1c | $2.53 \times 10^{-8}$ | -0.07 | -0.48 | -0.65 | -0.31 | mmol/mol | 5533 |
| TMC8 | missense<br>CADD $\geq 25$ | HbA1c | $3.01 \times 10^{-8}$ | 0.11 | 0.77 | 0.50 | 1.05 | mmol/mol | 2095 |
| HK1 | missense<br>CADD $\geq 25$ | HbA1c | $1.08 \times 10^{-7}$ | -0.16 | -1.08 | -1.48 | -0.68 | mmol/mol | 988 |
| SLC4A1 | missense<br>CADD $\geq 25$ | HbA1c | $1.76 \times 10^{-7}$ | -0.15 | -1.04 | -1.43 | -0.65 | mmol/mol | 1025 |
| TFR2 | missense<br>CADD $\geq 25$ | HbA1c | $4.24 \times 10^{-7}$ | -0.12 | -0.84 | -1.16 | -0.51 | mmol/mol | 1491 |
| CARHSP1 | missense<br>CADD $\geq 25$ | HbA1c | $6.78 \times 10^{-7}$ | -0.13 | -0.86 | -1.20 | -0.52 | mmol/mol | 1360 |

Given the large proportion of variants present in just a single individual, we used gene-based collapsing tests to look for associations with biomarkers of glycemic control and T2D diagnosis. We used two variant aggregation strategies; 1) pLOF variants with MAF ≤1% and 2) damaging missense variants with MAF ≤1% and performed burden testing in the unrelated White population (n=363,977) adjusting for age, sex and genetic ancestry via 12 principal components.

First, we tested genes for association with glucose and HbA1c levels. We required at least 10 variant carriers per gene to have measurements based on an examination of genomic inflation at different carrier thresholds (Supplementary Figure 1). Using a p-value threshold adjusted for the number of variant sets and phenotypes tested (p ≤ 7.82 × 10^-7^), four genes significantly associated with glucose levels: *GCK* pLOF (p = 1.56 × 10^-9^, 1.24 mmol/L increase), *GCK* damaging missense (p = 6.15 × 10^-11^, 0.61 mmol/L increase), *GIGYF1* pLOF (p = 4.42 × 10^-12^, 0.77 mmol/L increase) and *G6PC2* damaging missense variants (p = 4.62 × 10^-83^, 0.33 mmol/L decrease) (Figure 1, Table 1). The same variant sets also associated with HbA1c levels along with 27 additional sets including *HNF1A* pLOF (p = 2.14 × 10, 4.01 mmol/mol increase), *TNRC6B* pLOF (p = 2.36 × 10^-7^, 3.94 mmol/mol increase) and *PDX1* damaging missense variants (p = 2.54 × 10^-7^, 0.41 mmol/mol increase) (Figure 1, Table 1).

**Figure 1:**
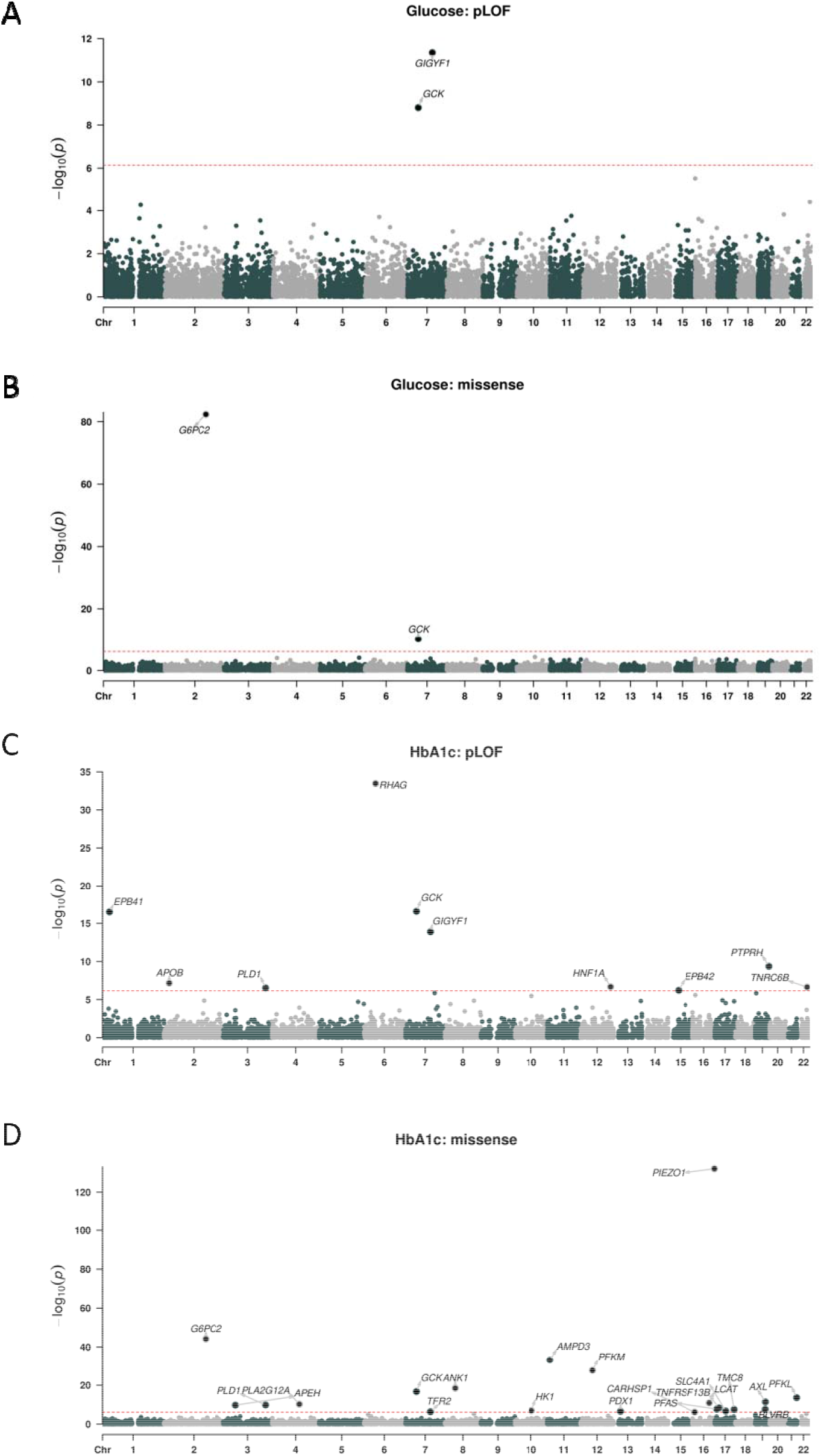
Gene-level associations with glucose and HbA1c levels. A) pLOF associations with glucose levels. B) Damaging missense variant (CADD score ≥ 25) associations with glucose levels. C) pLOF associations with HbA1c. D) Damaging missense variant associations with HbA1c levels. The red line indicates the threshold for significance, genes with significant associations are

We then tested aggregated pLOF and damaging missense variants for association with T2D diagnosis (n=24,695 cases). Using a p-value threshold adjusted for the number of variant sets tested (p ≤ 1.46 × 10^-6^), 6 variant sets significantly associated with T2D; pLOF variants in *GIGYF1, GCK, HNF1A* and *TNRC6B* and damaging missense variants in *GCK* and *PAM* (Figure 2, Table 2). As the time of available follow-up differs between England, Scotland, and Wales, we controlled for country of recruitment in the regression (see Methods). In addition, we confirmed that significant hits did not associate with country of recruitment (all p > 0.035) and that hits remained significant when only data from England were considered (Supplementary Table 2).

**Table 2:** Gene-level associations with T2D diagnosis. Association of pLOF or damaging missense variants (CADD score ≥ 25) aggregated per gene with T2D diagnosis. OR; odds ratio, CI; confidence interval.

| gene | Variant set | title | pvalue | OR | 95% CI - | 95% CI + | N cases | N carrier | N carrier cases | N expected |
| --- | --- | --- | --- | --- | --- | --- | --- | --- | --- | --- |
| GCK | pLOF | T2D | $2.96 \times 10^{-15}$ | 14.16 | 7.33 | 27.34 | 24695 | 40 | 19 | 2.71 |
| GIGYF1 | pLOF | T2D | $6.14 \times 10^{-11}$ | 4.15 | 2.71 | 6.37 | 24695 | 131 | 29 | 8.89 |
| HNF1A | pLOF | T2D | $1.23 \times 10^{-9}$ | 5.27 | 3.08 | 9 | 24695 | 73 | 20 | 4.95 |
| TNRC6B | pLOF | T2D | $2.00 \times 10^{-7}$ | 4.44 | 2.53 | 7.79 | 24695 | 71 | 17 | 4.82 |
| PAM | missense<br>CADD $\geq 25$ | T2D | $2.26 \times 10^{-12}$ | 1.31 | 1.21 | 1.41 | 24695 | 9357 | 801 | 634.85 |
| GCK | missense<br>CADD $\geq 25$ | T2D | $1.70 \times 10^{-8}$ | 2.96 | 2.03 | 4.32 | 24695 | 202 | 34 | 13.71 |

**Figure 2:**
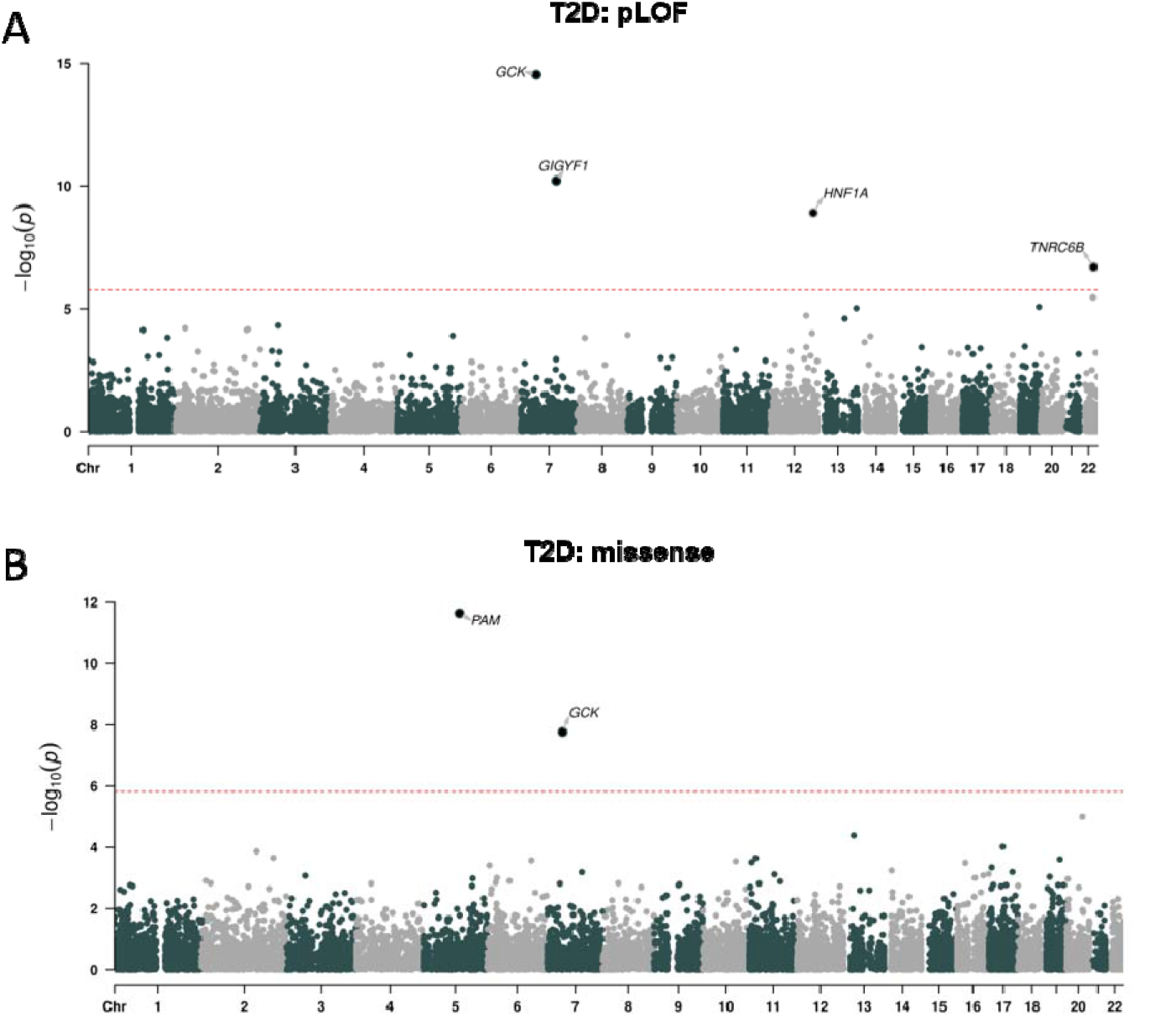
Gene-level associations with T2D. A) pLOF associations with T2D diagnosis. B) Damaging missense variant (CADD score ≥ 25) associations with T2D diagnosis. The red line indicates the threshold for significance, genes with significant associations are labeled.

### Identification of genes with a biological role in diabetes

Variants in two genes, *GCK* and *GIGYF1*, significantly associated with glucose, HbA1c and T2D diagnosis, strongly suggesting a biological role in diabetes; *GCK* is involved in Mendelian forms of diabetes while *GIGYF1* has not previously been implicated by genetics in the disease. Both *GCK* and *GIGYF1* are located on chromosome 7 but are 56Mb apart, strongly suggesting that these signals are independent; this independence was confirmed by conditional analysis (Supplementary Table 3). Two additional variant sets, *HNF1A* pLOF and *TNRC6B* pLOF, had genome-wide associations with both T2D diagnosis and HbA1c levels while *G6PC2* damaging missense associated with decreased levels of both glucose and HbA1c but not T2D diagnosis (Table 3).

**Table 3:** Genes and variant sets associated with multiple diabetes-related traits. Variant sets significant for at least one trait in our primary analysis that are also associated with additional diabetes traits (p ≤ 0.0016, 32 sets tested). Effect is shown in SD of transformed values or as an odds ratio (OR).

| gene | Variant set | Pvalue glucose | Effect glucose | Pvalue HbA1c | Effect Hba1c | Pvalue T2D | OR T2D |
| --- | --- | --- | --- | --- | --- | --- | --- |
| GCK | pLOF | $1.56 \times 10^{-9}$ | 0.999 | $2.64 \times 10^{-17}$ | 1.292 | $2.96 \times 10^{-15}$ | 14.16 |
| HNF1A | pLOF | 0.01 | 0.317 | $2.14 \times 10^{-7}$ | 0.592 | $1.23 \times 10^{-9}$ | 5.27 |
| GIGYF1 | pLOF | $4.42 \times 10^{-12}$ | 0.616 | $1.28 \times 10^{-14}$ | 0.639 | $6.14 \times 10^{-11}$ | 4.15 |
| GCK | missense<br>CADD $\geq 25$ | $6.15 \times 10^{-11}$ | 0.487 | $1.86 \times 10^{-17}$ | 0.565 | $1.70 \times 10^{-8}$ | 2.96 |
| PAM | missense<br>CADD $\geq 25$ | 0.92 | 0.001 | 0.009 | 0.026 | $2.26 \times 10^{-12}$ | 1.31 |
| TNRC6B | pLOF | $4.01 \times 10^{-5}$ | 0.507 | $2.36 \times 10^{-7}$ | 0.582 | $2.00 \times 10^{-7}$ | 4.44 |
| PDX1 | missense<br>CADD $\geq 25$ | 0.02 | 0.029 | $2.54 \times 10^{-7}$ | 0.060 | $3.99 \times 10^{-5}$ | 1.21 |
| PFAS | missense<br>CADD $\geq 25$ | 0.32 | 0.009 | $2.09 \times 10^{-8}$ | -0.048 | $4.43 \times 10^{-4}$ | 0.88 |
| G6PC2 | missense<br>CADD $\geq 25$ | $4.62 \times 10^{-83}$ | -0.266 | $6.71 \times 10^{-45}$ | -0.179 | 0.97 | 1.00 |

To see which other significant genes were likely to have a role in diabetes we looked at all variant sets with a significant glucose, HbA1c, or T2D association and examined whether they had associations with additional diabetes traits using a more permissive p-value threshold correcting for the number of variant sets tested (p ≤ 0.0016, 32 sets tested). Damaging missense variants in *PDX1* and *PFAS*, which had significant associations with HbA1c levels in our primary analysis, associated with T2D diagnosis using this threshold (Table 3 and Supplementary Table 4).

**Table 4:** PheWAS of *GIGYF1* pLOF – quantitative traits. Showing significant results for burden tests on quantitative traits (p ≤ 1.22 × 10^-4^). Effect is shown in standard deviations (SD) of transformed values. RH; right hand, LH; left hand.

| gene | variant set | title | pvalue | Effect (SD) | 95% CI - | 95% CI + | n carrier measured |
| --- | --- | --- | --- | --- | --- | --- | --- |
| GIGYF1 | pLOF | HbA1c | $1.28 \times 10^{-14}$ | 0.64 | 0.48 | 0.80 | 129 |
| GIGYF1 | pLOF | cholesterol | $2.44 \times 10^{-12}$ | -0.61 | -0.78 | -0.44 | 128 |
| GIGYF1 | pLOF | glucose | $4.42 \times 10^{-12}$ | 0.62 | 0.44 | 0.79 | 121 |
| GIGYF1 | pLOF | LDL cholesterol | $2.40 \times 10^{-10}$ | -0.56 | -0.73 | -0.38 | 128 |
| GIGYF1 | pLOF | apolipoprotein b | $2.52 \times 10^{-10}$ | -0.56 | -0.73 | -0.39 | 127 |
| GIGYF1 | pLOF | LH grip strength | $5.11 \times 10^{-10}$ | -0.37 | -0.49 | -0.25 | 131 |
| GIGYF1 | pLOF | RH grip strength | $1.01 \times 10^{-8}$ | -0.34 | -0.46 | -0.23 | 131 |
| GIGYF1 | pLOF | peak expiratory flow | $5.73 \times 10^{-8}$ | -0.41 | -0.56 | -0.26 | 114 |
| GIGYF1 | pLOF | cystatin c | $6.65 \times 10^{-6}$ | 0.36 | 0.20 | 0.51 | 128 |
| GIGYF1 | pLOF | mean corpuscular hemoglobin | $6.80 \times 10^{-6}$ | -0.38 | -0.55 | -0.22 | 128 |
| GIGYF1 | pLOF | HDL cholesterol | $1.53 \times 10^{-5}$ | -0.35 | -0.52 | -0.19 | 121 |
| GIGYF1 | pLOF | time to complete round (cognitive test) | $1.67 \times 10^{-5}$ | 0.35 | 0.19 | 0.51 | 129 |
| GIGYF1 | pLOF | waist circumference | $3.98 \times 10^{-5}$ | 0.32 | 0.16 | 0.47 | 130 |
| GIGYF1 | pLOF | total protein | $6.45 \times 10^{-5}$ | -0.36 | -0.53 | -0.18 | 121 |
| GIGYF1 | pLOF | apolipoprotein a | $6.88 \times 10^{-5}$ | -0.33 | -0.49 | -0.17 | 121 |

Many HbA1c associations appeared to be secondary to effects on red blood cells. 22 out of 31 variant sets associated with HbA1c did not show effects on glucose levels or T2D diagnosis (Supplementary Table 4) and were not implicated in Mendelian forms of diabetes. Out of these 22 variant sets, 12 were in genes implicated in Mendelian disorders affecting red blood cells (for example *EPB42* and *TFR2*; see Supplementary Table 5) and an additional five had highly significant associations with red blood cell traits in our data (p ≤ 7.82 × 10^-7^ ; Supplementary Table 6).

**Table 5:** PheWAS of *GIGYF1* pLOF – ICD10-coded diagnoses. Showing significant results for burden tests on ICD10 coded diagnoses with ≥ 500 cases and ≥ 1 expected case carrier (p ≤ 1.22 × 10^-4^). OR; odds ratio.

| gene | Variant set | title | pvalue | OR | 95% CI - | 95% CI + | N cases | N carrier cases | N expected |
| --- | --- | --- | --- | --- | --- | --- | --- | --- | --- |
| GIGYF1 | pLOF | E11 T2D | $6.14 \times 10^{-11}$ | 4.15 | 2.71 | 6.37 | 24695 | 29 | 8.89 |
| GIGYF1 | pLOF | E03 other hypothyroidism | $1.25 \times 10^{-9}$ | 4.53 | 2.78 | 7.38 | 19417 | 21 | 6.99 |
| GIGYF1 | pLOF | R55 syncope and collapse | $1.90 \times 10^{-6}$ | 3.75 | 2.18 | 6.47 | 12706 | 15 | 4.57 |
| GIGYF1 | pLOF | D50 iron deficiency anemia | $8.52 \times 10^{-6}$ | 3.56 | 2.04 | 6.23 | 12886 | 14 | 4.64 |
| GIGYF1 | pLOF | J43 emphysema | $1.99 \times 10^{-5}$ | 6.13 | 2.67 | 14.10 | 3015 | 6 | 1.09 |
| GIGYF1 | pLOF | N39 other disorders of urinary system | $7.32 \times 10^{-5}$ | 2.71 | 1.66 | 4.45 | 24581 | 19 | 8.85 |

**Table 6:** Common variant associations at the *GIGYF1* locus. Associations for the array-typed variant rs221783. For quantitative traits the effect is shown in standard deviations (beta) and for diagnoses as an odds ratio (OR). MAF; minor allele frequency.

| phenotype | chrom | Pos (hg19/hg38) | Ref (effect allele) | Alt | rsid | MAF | pvalue | Effect (beta/OR) | 95% CI - | 95% CI + |
| --- | --- | --- | --- | --- | --- | --- | --- | --- | --- | --- |
| glucose | 7 | 100292914/<br>100695291 | T | C | rs221783 | 11% | $1.82 \times 10^{-11}$ | -0.03 | -0.03 | -0.02 |
| HbA1c | 7 | 100292914/<br>100695291 | T | C | rs221783 | 11% | $3.58 \times 10^{-7}$ | -0.02 | -0.03 | -0.01 |
| Cholesterol | 7 | 100292914/<br>100695291 | T | C | rs221783 | 11% | $7.00 \times 10^{-12}$ | 0.03 | 0.02 | 0.03 |
| LDL | 7 | 100292914/<br>100695291 | T | C | rs221783 | 11% | $6.25 \times 10^{-10}$ | 0.02 | 0.02 | 0.03 |
| T2D | 7 | 100292914/<br>100695291 | T | C | rs221783 | 11% | 0.005 | 0.96 | 0.93 | 0.99 |
| Hypothyroidism | 7 | 100292914/<br>100695291 | T | C | rs221783 | 11% | $6.95 \times 10^{-7}$ | 0.92 | 0.88 | 0.95 |

We focused on the variant sets associated with multiple diabetes traits as these are strong candidates for regulating glucose homeostasis. The genes fall into three main groups; known MODY (maturity-onset diabetes of the young) genes (*GCK, HNF1A* and *PDX1*) [14], known genes reported in previous exome-wide analyses of glucose levels or T2D (*G6PC2* and *PAM*) [5, 15], and novel genes not previously implicated by genetics in diabetes (*GIGYF1, TNRC6B* and *PFAS*).

Because obesity is linked to the development of T2D, we adjusted for body mass index (BMI) in the burden tests and found that the association of variants in these genes with diabetes-related traits remained significant (Supplementary Tables 7 and 8).

Associations for rare variants can be susceptible to confounders such as population stratification and sample relatedness leading to false positives. Therefore, we used the generalized linear mixed model implemented by SAIGE-Gene which accounts for relatedness and adjusts for unbalanced case-control ratios [16] to verify association of our variant sets of interest with glucose, HbA1c, and T2D diagnosis. SAIGE-Gene was run in the White population including related individuals (n=398,574). Using the p-value thresholds previously employed, all associations were statistically significant using this method apart from the associations of *TNRC6B* pLOF with HbA1c (p = 6.85 × 10^-6^) and T2D diagnosis (p = 4.77 × 10^-5^) which were less significant (Supplementary Table 9).

To maximize power to detect associations for rare variants, our original analysis of glucose and HbA1c included individuals with a diabetes diagnosis. Associations for all variant sets of interest were at least nominally significant when such individuals were excluded from the analysis (Supplementary Table 10). For *GIGYF1* pLOF, there was still a substantial effect on glucose (p=2.95 × 10^-8^, effect = 0.53 SD) and HbA1c (p=8.29 × 10^-7^, effect = 0.43 SD) levels in carriers without a formal diabetes diagnosis.

### *GIGYF1* pLOF associations replicate using independent datasets

We sought to use independent measurements of glucose and HbA1c to verify the associations of interest seen in our primary analysis which used measurements taken as part of the UKBB assessment. To do this we extracted lab test values for glucose and HbA1c from primary care data, which is available for approximately half of the cohort, taking the mean measurement per individual. In gene-based burden tests all variant sets showed a direction of effect consistent with that seen in the primary analysis and 10 out of 12 of these were significant when correcting for the number of tests performed (p ≤ 0.004). This included the association of *GIGYF1* pLOF with glucose (p=2.10 × 10^-6^, effect = 0.65 SD) and HbA1c (p=1.19 × 10^-5^, effect = 0.74 SD) levels (Supplementary Figure 2 and Supplementary Table 11).

We then assessed whether rare variants in *GIGYF1* and the other novel genes associated with T2D replicated in an independent exome-sequencing cohort. Gene-based tests in European ancestry individuals from the Geisinger Health System (GHS; 25,846 T2D cases and 63,749 controls) confirmed the association of *GIGYF1* pLOF with T2D (p=0.01, OR=1.8). We did not replicate the association of *TNRC6B* pLOF with T2D. We also tested an expanded *PFAS* variant set (pLOF + deleterious missense) and did not detect an association with T2D (Supplementary Table 12). Notably variant set definitions varied somewhat from those used in our primary analysis (see Methods).

### Multiple variants contribute to associations with diabetes diagnosis and biomarkers

To examine whether specific variants were driving the associations with diabetes traits we conducted “leave-one-out” burden tests. The association of *PAM* missense variants with T2D diagnosis was driven entirely by a previously reported variant Ser539Trp (rs78408340; p = 0.43 when Ser539Trp is excluded). For all other variant sets, multiple variants contributed to the associations observed (Supplementary Figure 3). Notably, when singleton variants were excluded, half of the associations no longer reached significance including those for *GCK* pLOF and glucose (p = 0.0015 without singletons versus p = 1.56 x10^-9^) and *GIGYF1* pLOF and T2D (p = 2.9 × 10^-5^ without singletons versus p = 6.14x10^-11^) (Supplementary Table 13), demonstrating the power of including singletons in gene-based tests.

**Figure 3:**
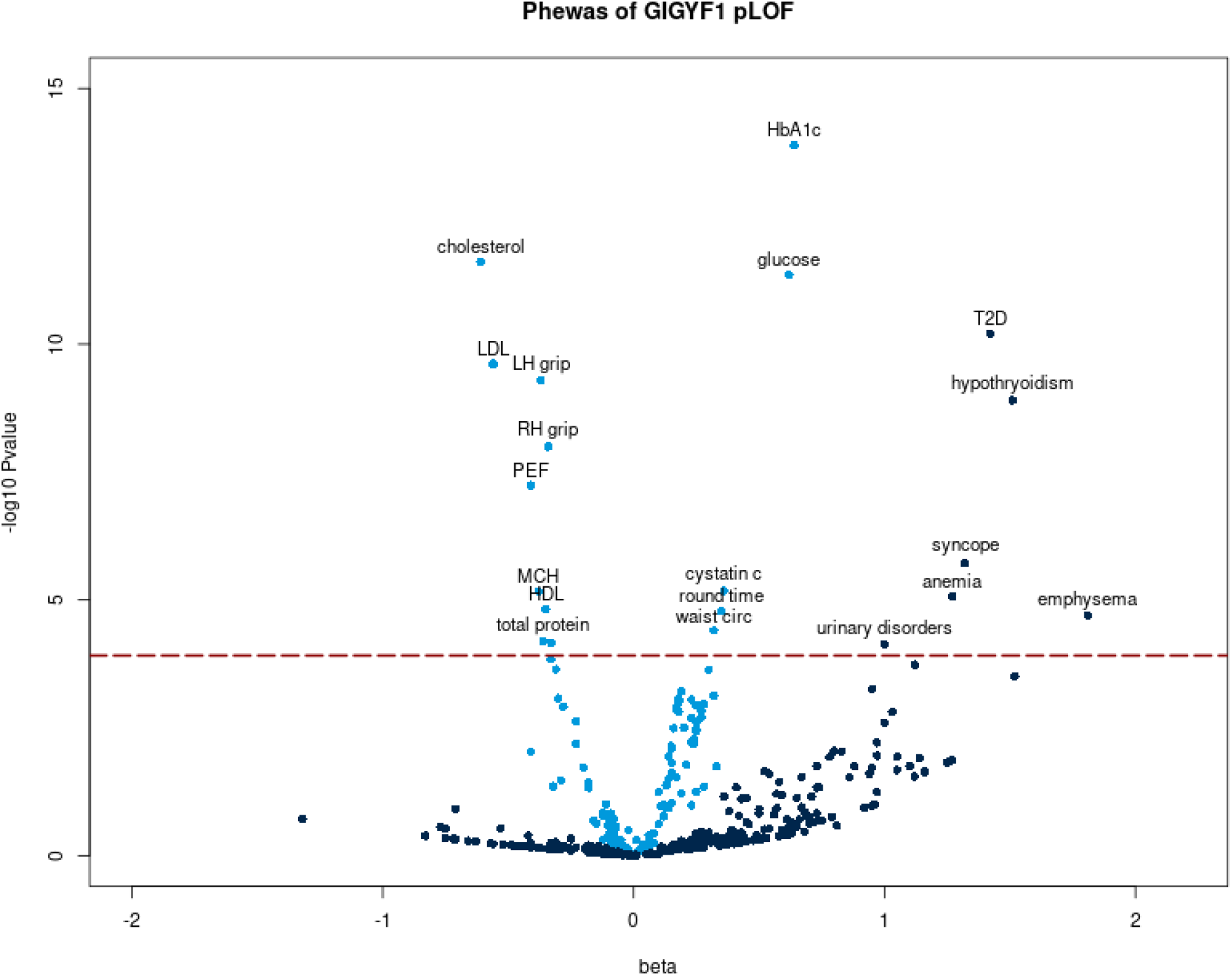
PheWAS of *GIGYF1*. pLOF The x-axis is the beta (effect size in standard deviations) for the association and the y-axis is -log10(p-value). Quantitative traits are colored light blue and ICD10 diagnoses colored dark blue. Phenome-wide significant associations are labeled. The dashed line indicates the p-value threshold for phenome-wide significance. Protein; total protein, RH grip; right hand grip strength, round time: time to complete round (cognitive test), LH grip; left hand grip strength, PEF; peak expiratory flow.

For the variants contributing to our novel discovered associations, *GIGYF1* pLOF, *TNRC6B* pLOF and *PFAS* damaging missense variants, we examined the quality scores, sequencing depth, transcripts affected and presence of contributing variants in gnomAD. We found that for *GIGYF1* and *PFAS* the variants contributing most to the associations had good quality scores and depth and were present in the non-Finnish European population in gnomAD. In contrast, *TNRC6B* is a highly constrained gene and the most common pLOF variant is not present in gnomAD. It is possible pLOF variants for constrained genes may not result in true loss of function (see Supplementary Note and Supplementary Figure 4). This observation along with the fact that the association of *TNRC6B* pLOF with T2D did not replicate in Geisinger Health System leads us to view this association with suspicion.

**Figure 4:**
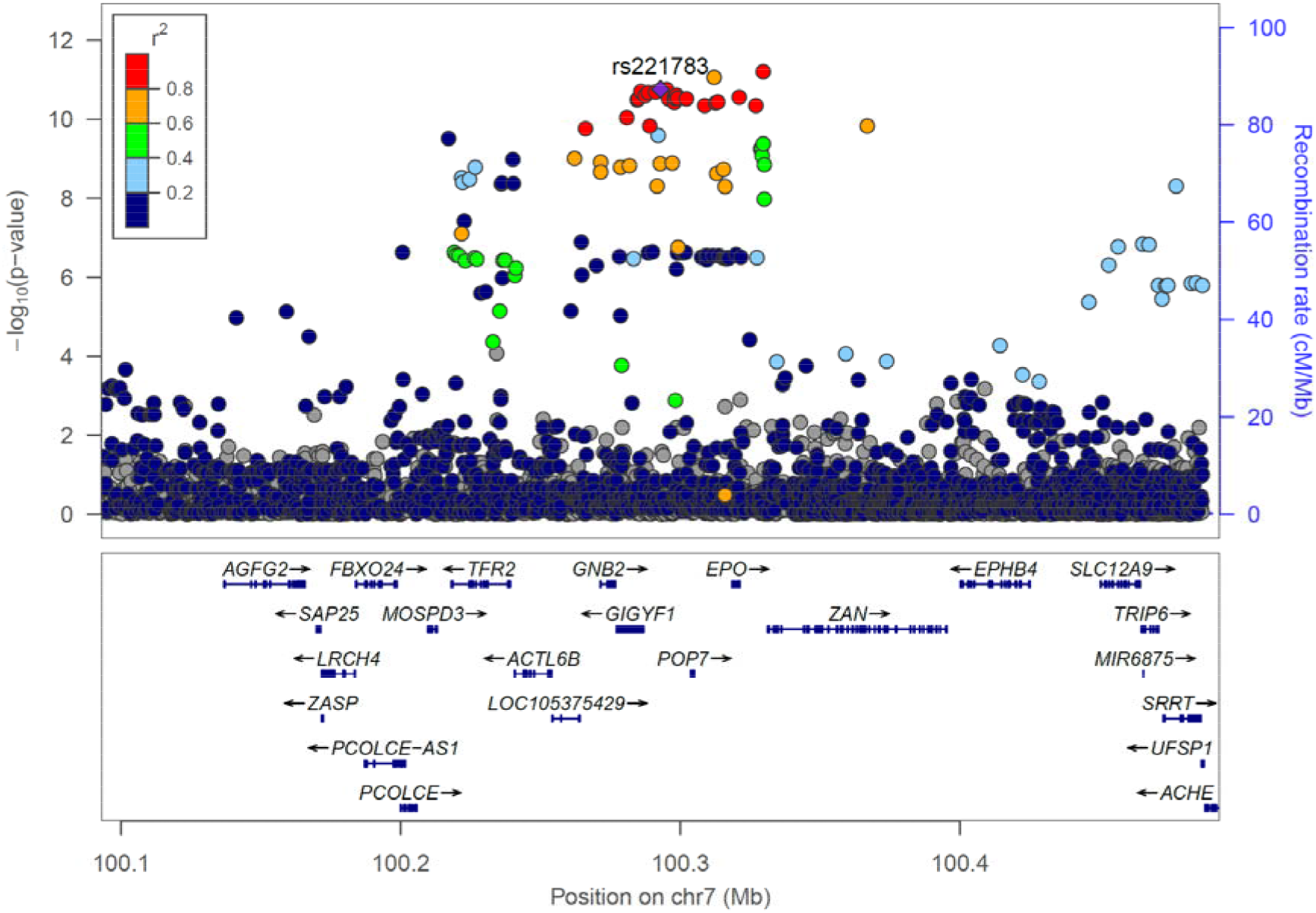
Locus plot of glucose associations at the *GIGYF1* locus. Association results for array genotyped and imputed variants are shown. The purple diamond represents the lead variant rs221783. Other variants are colored according to correlation (R^2^) with this marker (legend at top-left). The region displayed is chr7: 100092914-100492914. Genomic coordinates are for hg19.

### Replication of published gene-level associations with T2D and associations for T2D drug target genes

The association between predicted damaging variants in *PAM* and T2D diagnosis was previously reported in an exome-sequencing study performed by Flannick and colleagues [5]. We examined whether the other two significant genes in the study, *SLC30A8* and *MC4R*, associated with diabetes traits in our analysis. Both pLOF and damaging missense variants in *SLC30A8* associated with reduced levels of HbA1c and glucose and suggestively associated with decreased incidence of T2D diagnosis (Supplementary Table 14). Combining *SLC30A8* pLOF and missense variants resulted in more significant associations with glucose (p = 2.71 × 10^-6^), HbA1c (p = 8.64 × 10^-10^) and T2D diagnosis (p = 0.005) (Supplementary Table 14). There were no *MC4R* high confidence pLOF variants in our dataset and *MC4R* predicted damaging missense variants did not associate with diabetes-related traits in our study (all p > 0.19). We note that the *MC4R* Ile269Asn variant driving the association in Flannick and colleagues’ analysis is absent from our dataset, consistent with the fact that it is absent from European populations in gnomAD.

We also examined whether we detect associations for the 8 genes encoding T2D drug targets (*GLP1R, IGF1R, PPARG, INSR, SLC5A2, DPP4, KCNJ11, ABCC8*). Variant sets in three of these genes, *DPP4, GLP1R* and *KCNJ11* significantly associated with either T2D diagnosis or HbA1c levels (p < 0.003 correcting for 15 variant sets tested) and an additional 4 genes had a nominally significant association with T2D and/or HbA1c (Supplementary Figure 5 and Supplementary Table 15).

### PheWAS of *GIGYF1* pLOF reveals associations with cholesterol levels, hypothyroidism and complications of diabetes

The most significant novel associations were seen for *GIGYF1* pLOF which associated with increased glucose and HbA1c levels as well as increased incidence of T2D diagnosis. *GIGYF1* encodes a protein named for its binding to GRB10 (GRB10 interacting GYF protein 1), an adapter protein that has been shown to bind both the insulin and IGF-1 receptors. The association between *GIGYF1* pLOF and increased diabetes risk indicates that *GIGYF1* has a role in regulating insulin signaling and in protecting from diabetes. To give additional insight into the biological roles of GIGYF1 we performed a phenome-wide association study (PheWAS) testing *GIGYF1* pLOF for association with 142 quantitative traits and 262 ICD10-coded diagnoses. Based on the number of tests performed, the threshold for significance was p ≤ 1.22 × 10^-4^ (Figure 4).

*GIGYF1* pLOF strongly associated with decreased levels of total cholesterol (p=2.44 × 10^-12^, effect = -0.61 SD) which was, in large part, driven by LDL cholesterol (p = 2.40 × 10^-10^, effect = -0.56 SD) although an effect on HDL cholesterol was also observed (Table 4). To understand the extent to which this is influenced by the use of cholesterol-lowering medication in diabetics, we adjusted for medication use in the regression and performed a separate analysis excluding those on cholesterol-lowering medication. The association between *GIGYF1* pLOF and LDL cholesterol levels was significant in both analyses (Supplementary Table 16). *GIGYF1* pLOF also associated with decreased grip strength and decreased peak expiratory flow which may reflect changes in body size, muscle mass or general health in carriers [17, 18]. Notably, *GIGYF1* pLOF also associated with increased levels of the kidney injury biomarker cystatin c (p= 6.65 × 10^-6^, effect = 0.36 SD) and increased diagnosis of urinary system disorders (p = 7.32 × 10^-5^, OR = 2.71) which might suggest renal complications of diabetes in carriers (Table 4 and Table 5).

After diabetes, the most significant disease association of *GIGYF1* pLOF was with increased risk of hypothyroidism (p = 1.25 × 10^-9^, OR = 4.53). 21 out of the 131 *GIGYF1* pLOF carriers had a diagnosis of unspecified hypothyroidism and 7 of these also had a diagnosis of T2D. Given the autoimmune component in hypothyroidism and type 1 diabetes (T1D), we examined the association of *GIGYF1* pLOF with T1D diagnoses but did not detect a significant association (p = 0.1). *GIGYF1* pLOF significantly associated with increased risk of syncope and collapse (p = 1.92 × 10^-6^, OR = 3.75), possibly reflecting complications of diabetes or thyroid disorders (Table 5).

Other phenome-wide significant associations with quantitative traits included waist circumference, total protein and mean corpuscular hemoglobin as well increased time to complete a cognitive test (Table 4). To ensure that the association of *GIGYF1* pLOF with HbA1c was independent of effects on hemoglobin we adjusted for mean corpuscular hemoglobin level and verified that the association remained highly significant (p = 4.10 × 10^-12^). *GIGYF1* pLOF also associated with increased diagnosis of emphysema and anemia (Table 5).

### Common variants at *GIGYF1* associate with glucose, T2D and *GIGYF1* expression

Replication is a challenge for rare variant association studies. Despite the rarity of *GIGYF1* pLOF variants, we replicated the T2D association in an independent cohort. In addition, we looked for more common variants that could further implicate the *GIGYF1* locus in diabetes. We tested array genotyped and imputed variants at the *GIGYF1* locus for association with glucose levels in 294,042 unrelated White individuals with measurements available. We found a cluster of variants in a linkage disequilibrium block covering *GIGYF1* and *EPO* significantly associating with glucose levels (Figure 4). This signal is represented by rs221783, an intergenic variant whose minor T allele associated with decreased glucose (p = 1.8 × 10^-11^, effect = -0.03 SD,) and HbA1c (p = 3.6 × 10^-7^, effect = -0.02 SD,) levels as well as increased cholesterol (p = 7.0 × 10^-12^, effect = 0.03 SD,). This variant also associated with a decreased risk of T2D (p = 0.005, OR = 0.96) and hypothyroidism (p = 6.95 × 10^-7^, OR=0.92) (Table 6). rs221783 is the best eQTL (R^2^ > 0.8) for *GIGYF1* in several tissues including pancreas, adipose and thyroid [19] (Supplementary Table 17). In all tissues, the T allele associating with decreased glucose and decreased T2D risk associated with increased *GIGYF1* expression. Conditional analysis showed that the glucose and HbA1c associations of *GIGYF1* pLOF and rs221783 are independent of each other (Supplementary Table 18).

The association of rs221783 with glucose levels replicated in Biobank Japan (p = 1.7 × 10^-4^, effect = -0.05 SD for T allele) [20] whilst in FinnGen, rs221783 showed a nominal association with T2D diagnosis (p = 0.02, OR = 0.96 for T allele) (Supplementary Table 19). The association with thyroid disease has been replicated elsewhere [21].

The independent glucose and T2D associations at the *GIGYF1* locus and their replication in other datasets further support the hypothesis that decreasing GIGYF1 predisposes to diabetes while increasing GIGYF1 levels may protect from diabetes.

### Identification of causal genes at GWAS loci

Given the fact that the *GIGYF1* locus harbors both rare and common variants associated with T2D we examined whether our study points to the causal gene at additional GWAS loci. For 558 variants associated with T2D in a recent study by Vujkovic and colleagues [9] we tested whether either of the two closest genes associated with T2D or HbA1c levels in our study. Just nine genes close to these 558 variants significantly associated with T2D or HbA1c (p ≤ 2.41 × 10^-5^ adjusting for 2071 variant sets tested) - *ANK1, GCK, HNF1A, TNRC6B, SLC30A8, NF1, IRS2, CFTR* and *HNF4A* (Supplementary Figure 6 and Supplementary Table 20). Most of these genes are already known to be causal for T2D including *GCK, HNF1A, SLC30A8, IRS2* and *HNF4A*. Given that there is a common variant association with T2D at *TNRC6B* but conflicting results for *TNRC6B* pLOF in UKBB and GHS, further study of this locus may be warranted.

## Discussion

Our results highlight the power of whole exome sequencing to make novel discoveries relevant to human disease and to detect known associations of Mendelian disease genes. Gene-level aggregation and burden testing of rare pLOF and predicted damaging missense variants identified genes associating with diabetes and biomarkers of glycemic control. These included several genes not previously implicated in diabetes, *GIGYF1, TNRC6B* and *PFAS*, as well as *GCK, HNF1A* and *PDX1*, known MODY genes [14, 22-24]. We also identified *PAM* and *G6PC2*, genes highlighted by other rare-variant studies of T2D and glucose levels [5, 15]. Gene-level tests were needed to detect the majority of these associations owing to the rarity of the variants. For example, out of 363,977 individuals, just 40 carried a pLOF variant in *GCK* and 131 carried a pLOF variant in *GIGYF1*. In general, singleton variants contributed a large part of the signal arguing strongly, as others have done [4], for including such variants in gene-based collapsing tests.

Test statistic inflation can be a challenge when testing rare variants as statistical assumptions break down when the number of carriers expected to have the disease of interest is low [4, 25]. To avoid false positives in our analysis of diabetes, we initially examined associations with glucose and HbA1c because quantitative traits are less susceptible to inflation. All of the variant sets that associated with T2D also affected HbA1c and/or glucose levels giving us confidence in these associations. In addition, T2D associations for all genes, apart from *TNRC6B*, were significant (p ≤ 1.46 × 10^-6^) using the linear mixed model implemented by SAIGE-Gene which can be more robust when dealing with low numbers of variant carriers [16]. We also verified the majority of our associations with glucose and HbA1c levels, including those for *GIGYF1* pLOF, using independent measurements from primary care data. Additional confidence in our results comes from the fact that we identified genes known to be involved in Mendelian forms of diabetes and previously reported genes. In addition, a targeted analysis of the genes encoding T2D drug targets revealed HbA1c and/or T2D associations for variants in several of these genes. The lack of association for variants in some of these drug target genes may partly be due to a lack of statistical power. Several of these genes are constrained for pLOF variation and/or have small numbers of pLOF carriers in UKBB (for example, *PPARG* has just 16 pLOF carriers). However, for some of these genes such as *SLC5A2* (encoding SGLT2) we do not detect associations with diabetes traits despite good numbers of variant carriers.

We uncovered novel associations with T2D and biomarkers of glycemic control for aggregated variants in *GIGYF1, TNRC6B* and *PFAS* and attempted replication of these associations in exome-sequenced individuals from GHS. The association of *GIGYF1* pLOF with T2D replicated in this cohort but we did not replicate associations for *TNRC6B* and *PFAS* variants. There are differences between these two cohorts; UKBB is a population-based cohort with T2D diagnoses obtained from inpatient records while GHS is a health system-based cohort and includes both inpatient and outpatient diagnoses. There is a larger effect size for *GIGYF1* pLOF in UKBB compared to GHS which may be due to these differences in ascertainment. Differences in the definition of the variant sets tested especially for *PFAS* (see Methods) or the frequency of the relevant variants (for example, the frequency of *TNR6CB* pLOF is 0.01% in UKBB but 0.16% in GHS) may have contributed to the failure to replicate the *TNRC6B* and *PFAS* associations. Alternatively, this may suggest that the *TNRC6B* and *PFAS* associations are false positives.

We focused our analysis on understanding the consequences of *GIGYF1* pLOF as it strongly associated with glucose, HbA1c and T2D and the T2D association replicated in GHS. *GIGYF1* encodes a protein that was initially identified for its binding to the adapter protein GRB10 which negatively regulates both the insulin and IGF-1 receptors [26]. Transfection of cells with GRB10-binding fragments of GIGYF1 lead to greater activation of both the insulin and IGF-1 receptors [27]. This supports a hypothesis whereby GIGYF1 enhances insulin signaling by reducing the negative regulation of the insulin receptor by GRB10. When GIGYF1 is reduced, as is the case in individuals carrying pLOF variants, GRB10 presumably inhibits insulin signaling to a greater degree thereby reducing the action of insulin in its target tissues and leading to increased risk of T2D. However, the exact mechanistic details of these interactions remain to be determined. *GRB10* variants have also been reported to associate with T2D and glycemic traits although interpretation of these results is complicated by imprinting [28, 29]. *GIGYF1* is broadly expressed with high levels observed in endocrine tissues, pancreas and brain [19, 30]. GIGYF1 and the related protein GIGYF2 have also been implicated in translational repression [31] and translation-coupled mRNA decay [32] suggesting biological roles beyond regulation of insulin and IGF-1 receptor signaling.

PheWAS of *GIGYF1* pLOF revealed a strong association with decreased cholesterol levels reflecting altered energy homeostasis in carriers. An inverse relationship between glucose and cholesterol levels has been observed for variants in other genes [33]. We also observed several associations that could reflect complications of diabetes in *GIGYF1* pLOF carriers including increased cystatin c levels and increased diagnosis of urinary disorders, suggesting renal complications, as well as syncope and collapse which may be a side-effect of hyperglycemia and/or hypoglycemia in diabetics. Other associations may reflect poor health in carriers including decreased grip strength and decreased peak expiratory flow. *GIGYF1* pLOF also associated with decreased mean corpuscular hemoglobin levels and increased diagnosis of anemia as well as increased emphysema diagnosis. The biological basis for these associations is not clear. *GIGYF1* is highly expressed in lung [19, 30] although the emphysema association is driven by small numbers of individuals, so replication is required.

*GIGYF1* pLOF associated with a 4.5-fold increased risk of hypothyroidism and *GIGYF1* is highly expressed in thyroid [19, 30] consistent with a biological function in this tissue. IGF-1 and insulin have been implicated in the proliferation of thyroid cells which may, in part, explain the association with thyroid dysfunction [34-36]. An alternative possibility is that GIGYF1 contributes to thyroid function by affecting secretion of thyroid stimulating hormone in the anterior pituitary gland. Another explanation is that shared autoimmune mechanisms contribute to thyroid dysfunction and diabetes in pLOF carriers and that some of the carriers diagnosed with T2D have features of latent autoimmune diabetes in adults [37]. Damaging variants in *GIGYF1* have recently been implicated in conferring risk for developmental delay and autism spectrum disorders [38]. Consistent with this, we see an association of *GIGYF1* pLOF with increased time to complete a cognitive test. It may be that metabolic aberrations in carriers affect cognitive performance, that brain development is altered due to perturbation of IGF-1 signaling, or that other functions of GIGYF1 such as regulation of mRNA expression and decay are responsible for cognitive phenotypes.

In addition to replicating the association of *GIGYF1* pLOF with T2D in an independent cohort we also used common genetic variants to further investigate the role of the *GIGYF1* locus in diabetes. Non-coding variants at the *GIGYF1* locus associated with glucose levels and T2D, and we replicated these findings in independent datasets. These variants associated with increased *GIGYF1* expression but a lower risk of T2D. This direction of effect is consistent with what we see for the pLOF variants – reduced levels of *GIGYF1* increases diabetes risk but increased levels of *GIGYF1* are protective.

We observed an intersection of rare and common variant associations at *GIGYF1* as well as at MODY genes such as *GCK, HNF1A* and *HNF4A*. However, in general, our gene-level analysis of rare variants did not identify many additional causal genes at GWAS loci; out of 558 variants associated with T2D [9] just nine had rare variant associations at a nearby gene.

We assessed the impact of pLOF and predicted damaging missense variants in approximately 17,000 genes on glycemic traits and uncovered a hitherto unappreciated role for *GIGYF1* in regulating blood sugar and protecting from T2D. By highlighting the importance of GIGYF1 and GRB adapter proteins in modulating insulin signaling this finding may lead to new therapeutic approaches for the treatment of diabetes. Discoveries such as this are only possible by combining health-related data with the sequencing of rare variants on a biobank scale.

## Methods

### The UK Biobank resource and data access

The UK Biobank (UKBB) recruited ∼500,000 participants in England, Wales, and Scotland between 2006 and 2010 [39]. Written informed consent was obtained from all participants. Phenotypic data available includes age, sex, biomarker data and self-reported diseases collected at the time of baseline assessment as well as disease diagnoses from inpatient hospital stays, the cancer registry and death records obtained through the NHS. Approximately half of the participants also have diagnoses from primary care available. Array genotypes are available for nearly all participants and exome sequencing data is available for 454,787 participants. The data used in this study were obtained from the UKBB through application 26041.

### Population definition and PC calculation for subjects with exome data

Subject quality control was performed by Regeneron Genetics Center (RGC) and removed subjects with evidence of contamination, unresolved duplications, sex discrepancies and discordance between exome sequencing and genotyping data. Genetic relationships between participants were determined by RGC using the PRIMUS program [40]. For the unrelated subset all first- and second-degrees relatives and some third-degree relatives were excluded.

Populations were defined through a combination of self-reported ethnicity and genetic principal components. We selected the unrelated individuals who identify as White (Field 21000) and ran an initial principal component analysis (PCA) on high quality common variants using eigenstrat [41]. SNPs were filtered for missingness across individuals < 2%, MAF > 1%, regions of known long range LD [42], and pruned to independent markers with pairwise LD < 0.1. We then projected the principal components (PCs) onto related individuals and removed all individuals +/-3 standard deviations from the mean of PCs 1-6. A final PC estimation was performed in eigenstrat [41] using unrelated subjects. We then projected related individuals onto the PCs.

### Exome sequencing and variant calling

DNA was extracted from whole blood and was sequenced by the RGC as described elsewhere [43]. Briefly, the xGen exome capture was used and reads were sequenced using the Illumina NovaSeq 6000 platform. Reads were aligned to the GRCh38 reference genome using BWA-mem [44]. Duplicate reads were identified and excluded using the Picard MarkDuplicates tool (Broad Institute). Variant calling of SNVs and indels was done using the WeCall variant caller (Genomics Plc.) to produce a GVCF for each subject. GVCFs were combined to using the GLnexus joint calling tool [45]. Post-variant calling filtering was applied using the Goldilocks pipeline [43]. Variants were annotated using the Ensembl Variant Effect Predictor v95 [46] which includes a LOFTEE plug-in to identify high confidence (HC) pLOF variants [13]. Combined Annotation Dependent Depletion (CADD) scores were generated using the Whole Genome Sequence Annotator (WGSA) AMI version 0.8.

### Phenotype definitions

Blood biochemistry values were obtained for glucose (Field 30740) and HbA1c (Field 30750) from UKBB and inverse rank normalized using the RNOmni R package [47], resulting in an approximately normal distribution.

For disease diagnoses, ICD10 codes were obtained from inpatient hospital diagnoses (Field 41270), causes of death (Field 40001 and 40002) and the cancer registry (Field 40006) from UKBB. Diagnoses also included additional hospital episode statistics (HESIN) and death registry data made available by UKBB in July 2020. T2D was defined as ICD10 E11. For the purposes of excluding diagnosed diabetics from the glucose and HbA1c analysis we defined diabetes as ICD10 codes E10-E14 which includes both T1D and T2D diagnoses.

For phenome-wide analyses, a selection of quantitative traits was obtained from other fields, encompassing anthropometric measurements, blood counts, as well as blood and urine biochemistry. Beyond these measurements, we selected additional quantitative traits found to be heritable (h^2^ significance flagged as at least “nominal” with a confidence level flagged as “medium” or “high”) by the Neale lab [25], using PHESANT to transform values to quantitative traits when necessary as they describe. These included the results of cognitive tests. All quantitative traits were inverse rank normalized using the RNOmni R package. [47]. For burden testing, we required at least 10 carriers to have measurements. We also tested associations with ICD10-coded diagnoses (using 3 character codes) that had more than 500 cases in the White subset of participants with exome data and at least one expected case carrier based on variant frequency and disease prevalence.

Glucose and HbA1c values were also extracted from primary care data available for about half of the cohort using the following read codes. Glucose: read 2 codes 44U..,44g.., 44g1.,44TJ.,44f..,44TK.,44f1.,44g0.,44f0. and read 3 codes XM0ly, X772z, XE2mq; HbA1c: read 2 codes 42W5., 44TB., 66Ae0, 44TC., 42W4. and read 3 codes XaPbt, X772q, XaWP9, XaBLm, XaERp.

Values were converted to IFCC units where necessary. Aberrantly high (≥ 45 mmol/L for glucose, ≥ 300 mmol/mol for HbA1c) and extremely low values (≤ 0.6 mmol/L for glucose, ≤ 10 mmol/mol for HbA1c) were excluded. The mean measurement per individual was then taken and inverse rank normalized prior to association testing. The mean age at measurement was also extracted and used as a covariate in the regression.

Individuals taking cholesterol-lowering medication were identified using self-reported medications recorded at their UKBB interview (Field 20003) and whether cholesterol-lowering medications were recorded using the touchscreen questionnaire (Fields 6177 and 6153).

### Gene-based association testing

For gene-based tests, autosomal rare pLOF variants were identified as follows; LOFTEE high confidence LOFs, MAF ≤ 1%, missingness across individuals ≤ 2%, HWE p-value ≥ 10^-10^. Predicted damaging missense variants were defined as missense variants with a CADD PHRED-scaled score ≥ 25, MAF ≤ 1%, missingness across individuals ≤ 2%, HWE p-value ≥ 10^-10^. Only genes with more than one pLOF variant or damaging missense variant were tested.

Burden testing was performed unrelated White subset using glm in R, using a gaussian model for quantitative traits and a binomial model for case-control analyses. Genotype was coded as 0 (no variant) or 1 (any number of variants). We adjusted for age, sex and the first 12 PCs of genetic ancestry in the regression. Additionally, when testing for association with disease diagnoses, we included country of recruitment as a covariate as the time of available follow-up differs between England, Scotland and Wales. Recruitment country was defined using the location of the relevant UKBB recruitment center (Field 54). Associations were later confirmed using just participants recruited in England. For case-control analyses we only ran tests where there was at least one expected case carrier based on variant frequency and disease prevalence. For quantitative traits we required at least 10 carriers to have measurements.

For glucose and HbA1c, to convert effect sizes from normalized values back to measured units, the estimates from the regression were multiplied by the standard deviation of these traits in the entire cohort.

SAIGE-Gene was run using the SAIGE R package (v0.36.5) [48] using settings recommend by the developers and related individuals were included.

T2D drug targets were defined according to Flannick et al. [5].

Manhattan plots were created using the R Package CMplot (https://github.com/YinLiLin/R-CMplot).

### Array association testing

Genotypes were obtained through array typing and imputation as described previously [49]. Population definition and PC estimation for individuals with array data was performed as previously described [50]. We tested all variants with imputation quality score (info) ≥ 0.8 and minor allele frequency (MAF) ≥ 0.1% in a 200Mb region around *GIGYF1* for association with glucose, HbA1c, T2D and hypothyroidism. Association analyses were performed using an additive model in PLINK adjusting for age at recruitment to UKBB, sex and the first 12 PCs of genetic ancestry. We also adjusted for country of recruitment where appropriate. The most significant variant with info > 0.95 was selected as the lead variant at the locus.

We replicated the association of rs221783 with glucose using available summary statistics for Biobank Japan for the trait “blood sugar” (http://jenger.riken.jp/en/result) [20]. We replicated the association of this variant with T2D diagnosis using summary statistics from FinnGen release 3 for the phenotype “E4_DM2” (https://www.finngen.fi/en/access_results). The effect allele in these datasets was the alternate allele “C”. For consistency with the UKBB associations we have shown the effect for the “T” allele.

Meta-analysis of the UKBB and replication dataset association results was performed with the METAL software package using the classical method [51].

Region plots were created using LocusZoom [52]. LD calculations were performed in the White population for array variants in a 500kb sliding window as follows; we extracted genotypes with info > 0.9, rounded them to whole numbers, mean-imputed missing genotypes and used the R “cor” function to compute R which was then squared to get an R^2^ value.

### Gene expression and eQTL analysis

The expression of *GIGYF1* in various tissues was assessed using the GTEx portal (accessed 08/04/2020) [19] and Human Protein Atlas (http://www.proteinatlas.org) [30]. eQTL data for rs221783 was obtained from GTEx v8. For each tissue of interest, the best eQTL for *GIGYF1* was identified (GTEx v8 “eGene”). R^2^ for rs221783 and the best *GIGYF1* eQTL was calculated as described above.

### Replication analysis in GHS

The GHS MyCode Community Health Initiative study is a health system-based cohort and has been described previously [53]. A subset of participants sequenced as part of the GHS-Regeneron Genetics Center DiscovEHR partnership were included in this study. T2D status was defined based on meeting at least one of the following criteria: (1) clinical encounters due to or problem-lists diagnosis code for type 2 diabetes (ICD-10 code E11), or (2) HbA1c greater than 6.5%, or (3) use of diabetic oral hypoglycemic medicine. Controls were participants who did not meet any of the criteria for case definition. Individuals were excluded from the analysis if they had clinical encounters due to or problem-lists diagnosis code for type 1 diabetes (ICD-10 code E10), or if they were treated with insulin but not with oral hypoglycemic medicines.

Exome-sequencing, variant calling, quality control and gene-based tests were performed as previously described [54]. Variant sets tested were pLOF variants (*GIGYF1* and *TNRC6B*) or pLOF plus missense variants predicted to be deleterious by 5/5 algorithms (*PFAS*) with MAF < 1%. The following variants were classified as pLOF variants: frameshift-causing indels, variants affecting splice acceptor and donor sites, variants leading to stop gain, stop loss and start loss. The five missense deleterious algorithms used were SIFT [55], PolyPhen2 (HDIV), PolyPhen2 (HVAR) [56], LRT [57], and MutationTaster [58]. Association testing was performed in the European ancestry population using the Firth logistic regression test implemented in REGENIE [59] as previously described [54].

### Identification of potential causal genes at GWAS loci

For 558 variants identified as associating with T2D [9] we mapped the two closest protein coding genes using bedtools. This resulted in 1118 genes for which we had tested 2071 variant sets (pLOF and/or damaging missense) in our primary analysis. Genes with p < 2.41 × 10^-5^ (correcting for 2071 variant sets tested) for HbA1c or T2D were considered significant.

### Ethics Statement

The UK Biobank resource is an approved Research Tissue Bank and is registered with the Human Tissue Authority, which means that researchers who wish to use it do not need to seek separate ethics approval (unless re-contact with participants is required). Research in GHS was approved by the GHS IRB, approval number 2006-0258. Written informed consent was obtained from all participants in UKBB and GHS.

## Data Availability

The data used in this study were obtained from the UKBB through application 26041. All phenotypic data and array genotypes are accessible through application to UK Biobank. Currently, exome sequencing data for ∼200,000 participants is available; the remainder of the exome data is scheduled for public release in 2021.

## Data availability

All phenotypic data and array genotypes used in this study are accessible through application to UKBB. Currently, exome sequencing data for ∼200,000 participants is available [38]; the remainder of the exome data used is scheduled for public release in 2021. Summary statistics for gene-level tests will be made available upon publication.

## Author Contributions

A.D, L.W., M.P., A.F.C. and P.N. performed computational analyses; P.A., L.L. A.B. and RGC performed replication analysis in GHS; A.D. wrote the manuscript. All authors interpreted results and edited the manuscript.

## Acknowledgements

This research has been conducted using the UK Biobank Resource (Project 26041). We would like to thank the participants and researchers of UK Biobank for creating an open-access resource. We thank the UK Biobank Exome Sequencing Consortium and UK Biobank for facilitating exome sequencing of participants. We also thank the participants of the GHS MyCode initiative as well as participants and investigators of the FinnGen study and Biobank Japan. We thank Mark McCarthy and Anna Gloyn for comments on the manuscript. Data management and analytics were performed using the REVEAL/SciDB translational analytics platform from Paradigm4.

