## Supplemental information for "Gene-level analysis of rare variants in 363,977 whole exome sequences identifies an association of *GIGYF1* loss of function with type 2 diabetes"

* For full author and contribution list see Supplementary Information.

### Supplementary Information

### Supplementary Figures

#### Supplementary Figure 1: QQ plots for gene-based burden tests.

QQ plots stratified by the number of carriers measured are shown for A) glucose and B) HbA1c. QQ plots stratified by the minimum number of expected case carriers is shown for C) T2D diagnosis. The genomic inflation factor lambda at different thresholds is displayed.

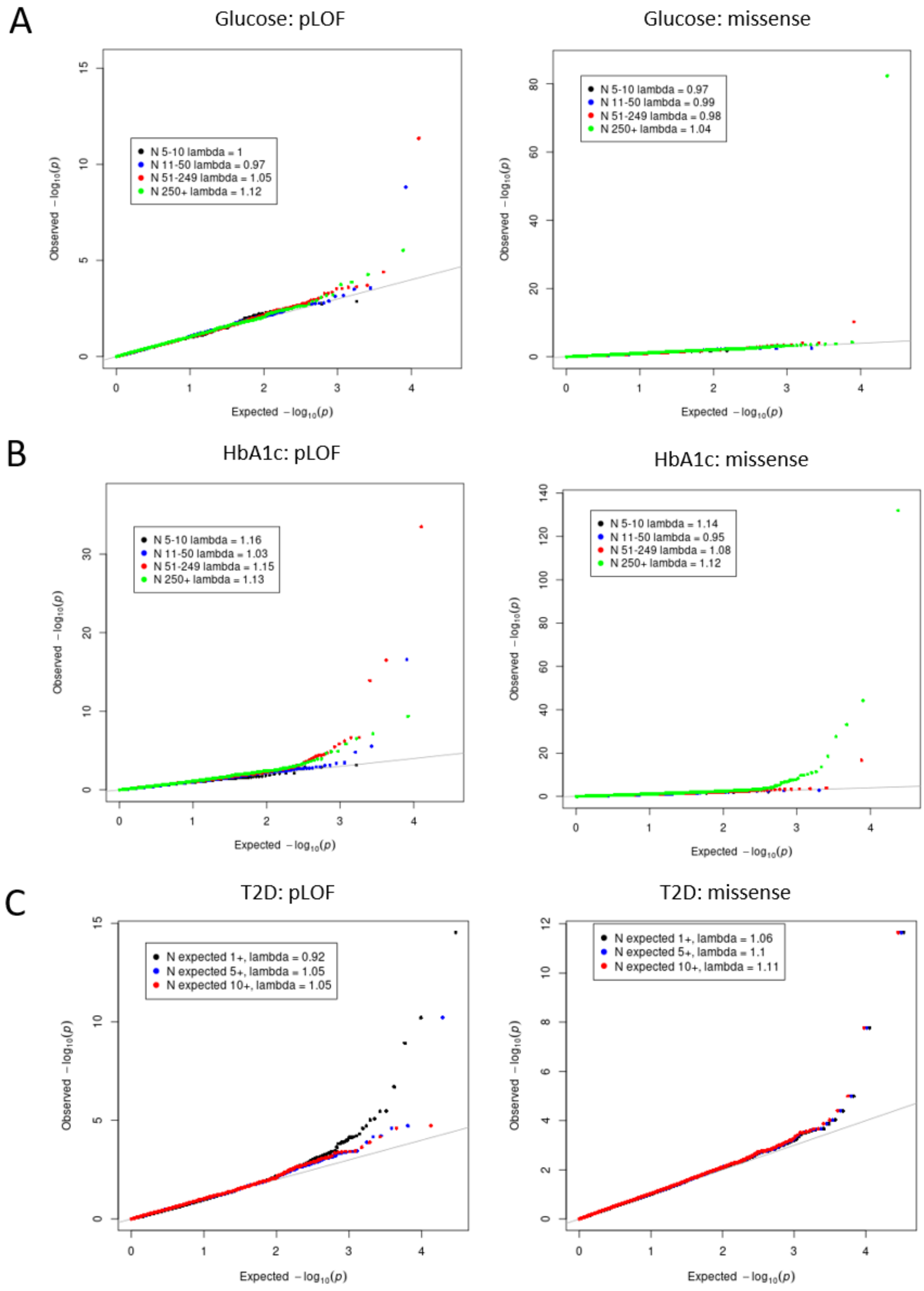
A
Glucose: pLOF
N 5-10 lambda = 1
N 11-50 lambda = 0.97
N 51-249 lambda = 1.05
N 250+ lambda = 1.12
Glucose: missense
N 5-10 lambda = 0.97
N 11-50 lambda = 0.99
N 51-249 lambda = 0.98
N 250+ lambda = 1.04
B
HbA1c: pLOF
N 5-10 lambda = 1.16
N 11-50 lambda = 1.03
N 51-249 lambda = 1.15
N 250+ lambda = 1.13
HbA1c: missense
N 5-10 lambda = 1.14
N 11-50 lambda = 0.95
N 51-249 lambda = 1.08
N 250+ lambda = 1.12
C
T2D: pLOF
N expected 1+, lambda = 0.92
N expected 5+, lambda = 1.05
N expected 10+, lambda = 1.05
T2D: missense
N expected 1+, lambda = 1.06
N expected 5+, lambda = 1.1
N expected 10+, lambda = 1.11
Observed $-\log_{10}(p)$
Expected $-\log_{10}(p)$

#### Supplementary Figure 2: Associations with glucose and HbA1c levels from primary care data

Forest plot showing associations with glucose and HbA1c levels as measured in primary care (GP) data. Effect size in standard deviations is shown along with the 95% confidence interval. The threshold for significance is $p \leq 0.004$ correcting for the number of genes and variant sets tested.

| Gene | Variant set | biomarker | p-value |
|---|---|---|---|
| GIGYF1 | pLOF | Glucose GP | 2.10e-06 |
| GCK | pLOF | Glucose GP | 1.41e-08 |
| GCK | missense | Glucose GP | 2.53e-03 |
| G6PC2 | missense | Glucose GP | 1.60e-20 |
| G6PC2 | missense | HbA1c GP | 4.14e-06 |
| GCK | pLOF | HbA1c GP | 3.90e-06 |
| GCK | missense | HbA1c GP | 6.25e-02 |
| GIGYF1 | pLOF | HbA1c GP | 1.19e-05 |
| HNF1A | pLOF | HbA1c GP | 5.68e-04 |
| PDX1 | missense | HbA1c GP | 3.41e-03 |
| PFAS | missense | HbA1c GP | 1.27e-03 |
| TNRC6B | pLOF | HbA1c GP | 1.15e-02 |

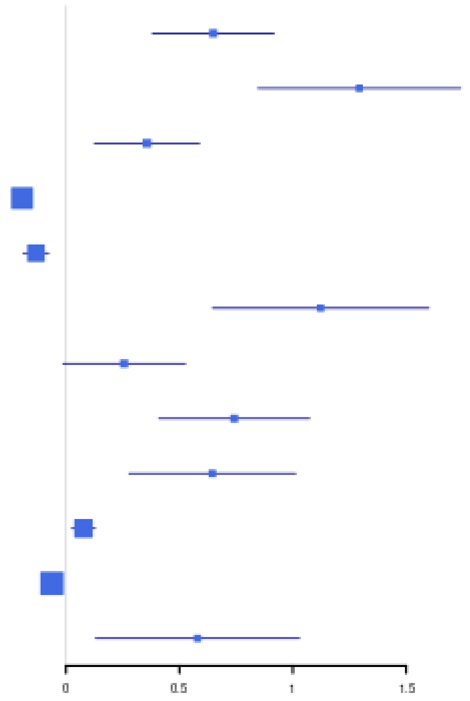

#### Supplementary Figure 3: Leave one variant out analysis

Burden tests were run for significant variant sets of interest leaving out one variant at a time. Negative log10 of the p-value for each test is shown relative to the genomic position of the excluded variant. A) Glucose associations B) HbA1c associations C) T2D associations. pLOF variant sets are labeled as “lof.hc” and damaging missense variant sets as “cadd.25”.

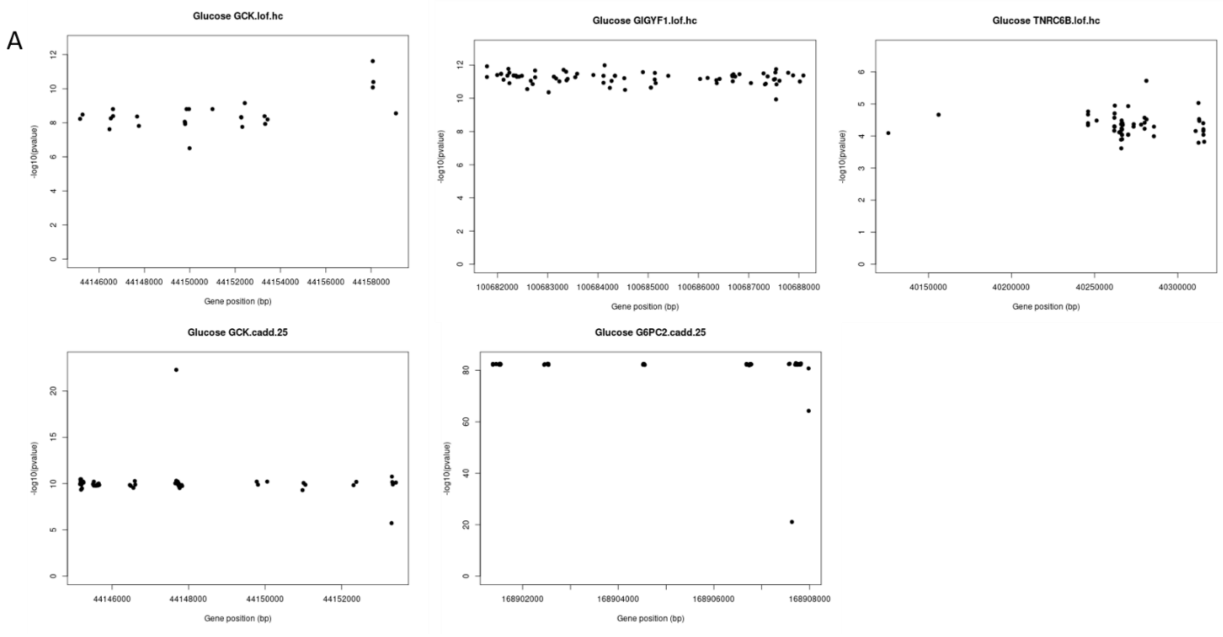

B

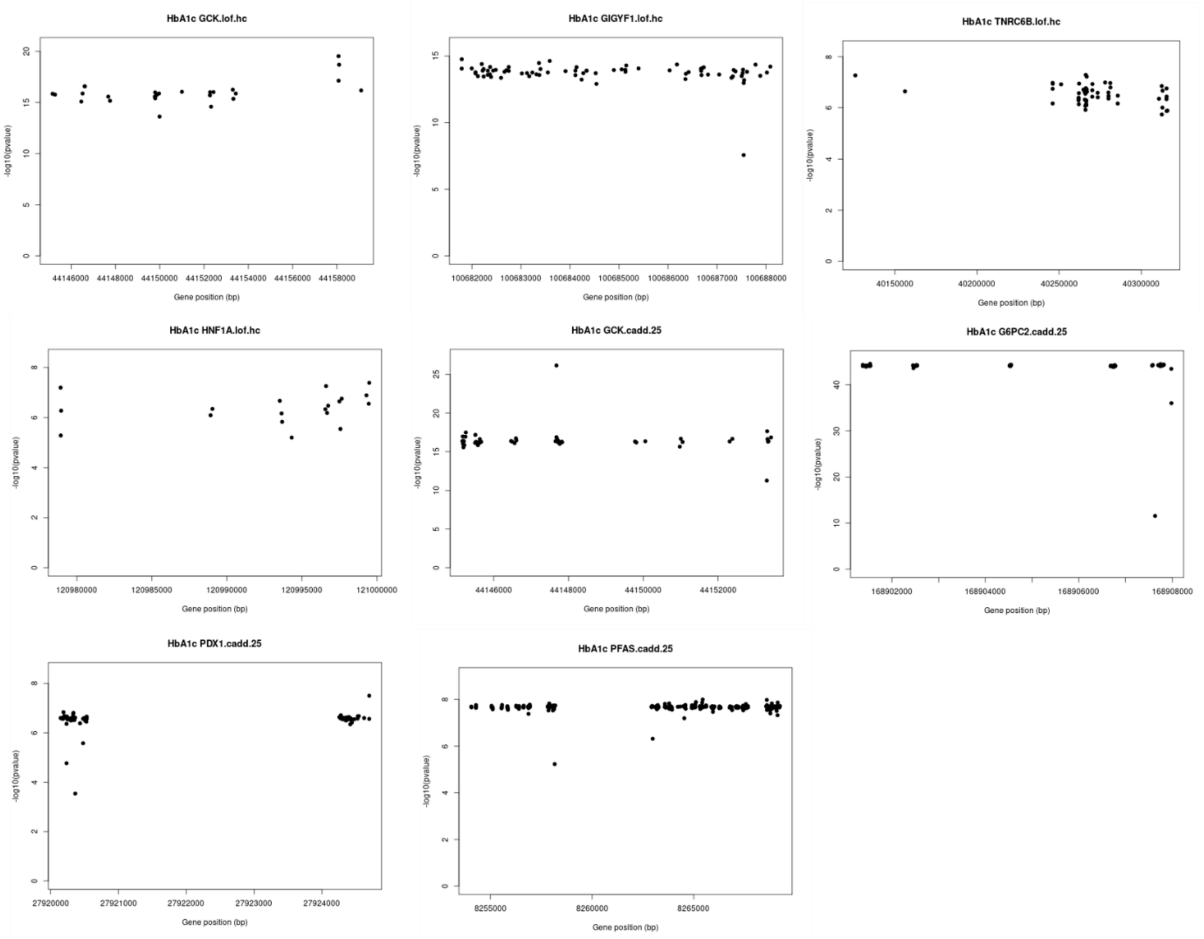
HbA1c GCK.lof.hc
HbA1c GIGYF1.lof.hc
HbA1c TNRC6B.lof.hc
HbA1c HNF1A.lof.hc
HbA1c GCK.cadd.25
HbA1c G6PC2.cadd.25
HbA1c PDX1.cadd.25
HbA1c PFAS.cadd.25
-log10(pvalue)
Gene position (bp)

C

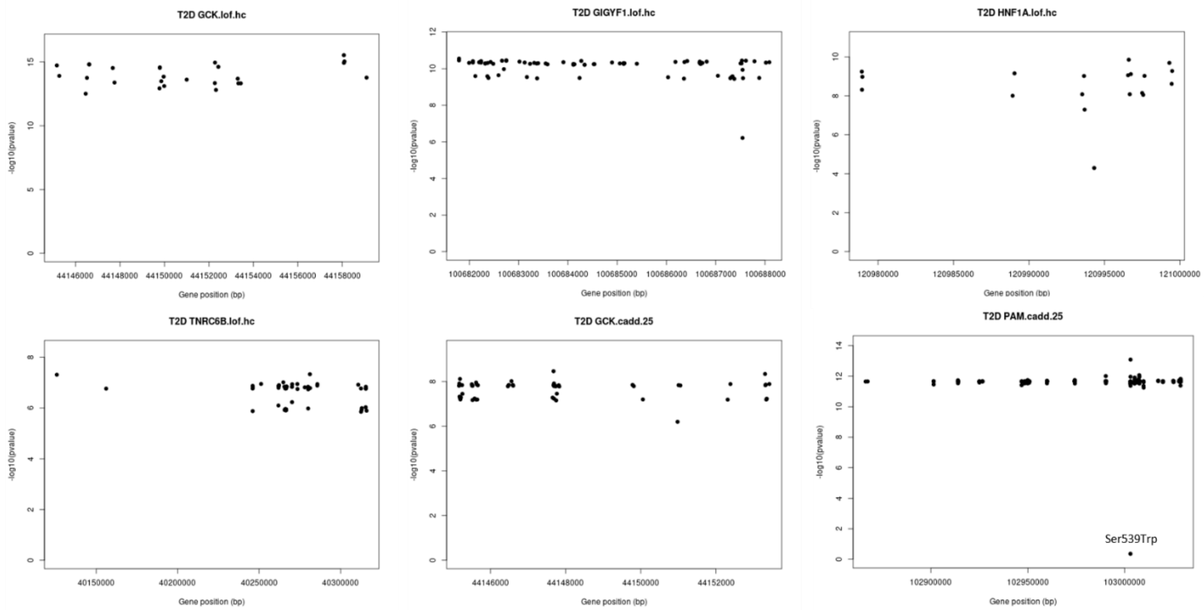
T2D GCK.lof.hc
T2D GIGYF1.lof.hc
T2D HNF1A.lof.hc
T2D TNRC6B.lof.hc
T2D GCK.cadd.25
T2D PAM.cadd.25
Ser539Trp
-log10(pvalue)
Gene position (bp)

#### Supplementary Figure 4: Location of variants of interest in *GIGYF1, PFAS* and *TNRC6B*

The locations of individual variants making up the variant sets are plotted with respect to Ensembl transcripts. Selected variants are shown as a separate track. A) *GIGYF1* pLOF variants, 7:100687545:CA:C shown separately. B) *PFAS* damaging missense variants, 17:8258163:G:A (Glu434Lys) and 17:8262979:G:A (Ala466Thr) are shown separately. C) *TNRC6B* pLOF variants, 22:40270205:G:T and 22:40281161:G:T shown separately.

A

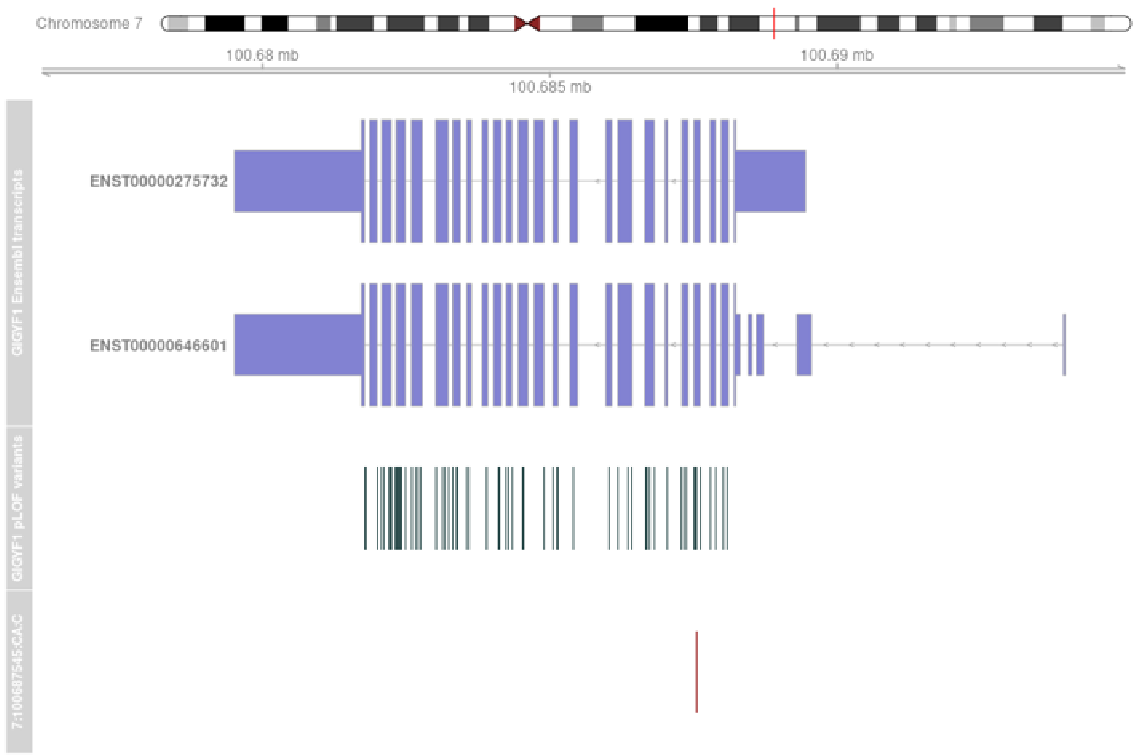
Chromosome 7
100.68 mb
100.69 mb
100.685 mb
GIGYF1 Ensembl transcripts
ENST00000275732
ENST00000646601
GIGYF1 pLOF variants
7:100687545:CA:C

B

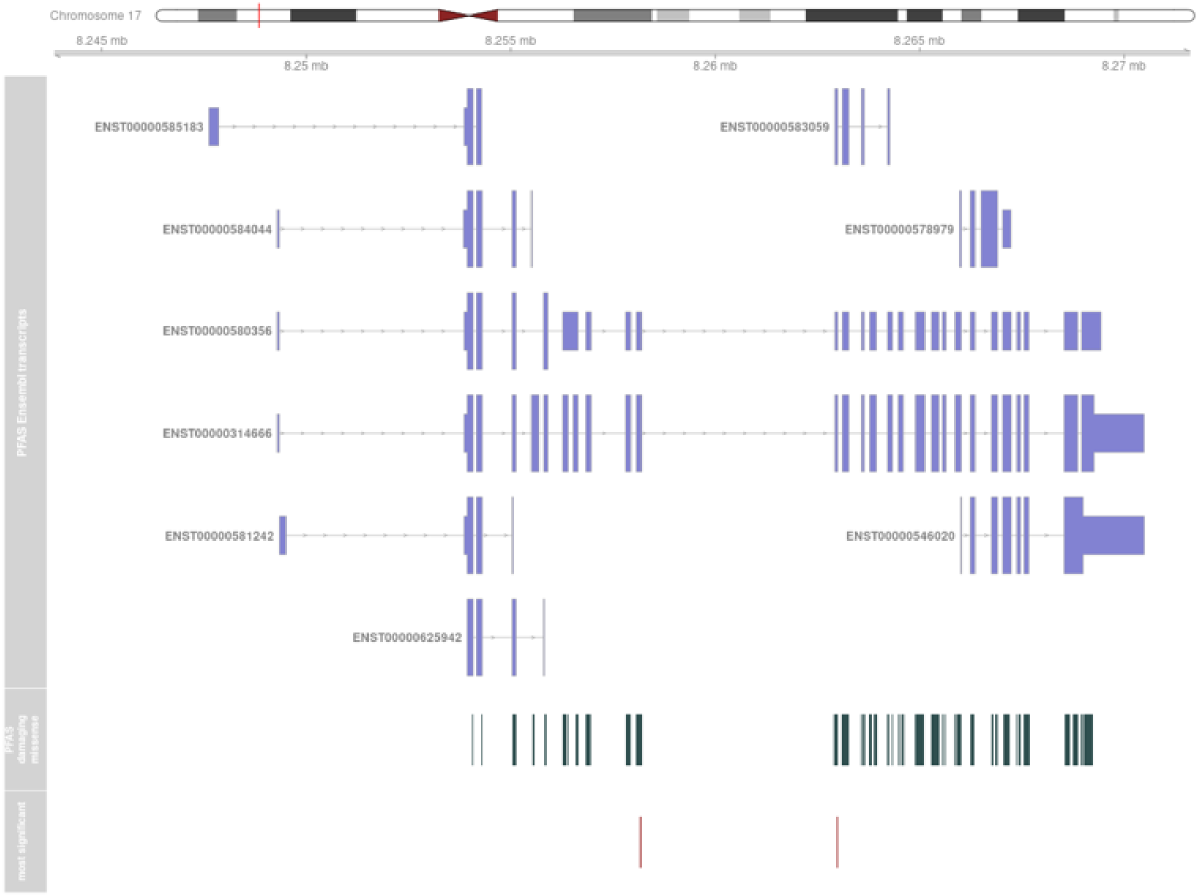
Chromosome 17
8.245 mb
8.255 mb
8.265 mb
8.25 mb
8.26 mb
8.27 mb
PFAS Ensembl transcripts
ENST00000585183
ENST00000583059
ENST00000584044
ENST00000578979
ENST00000580356
ENST00000314666
ENST00000581242
ENST00000546020
ENST00000625942
PFAS damaging missense
most significant

C

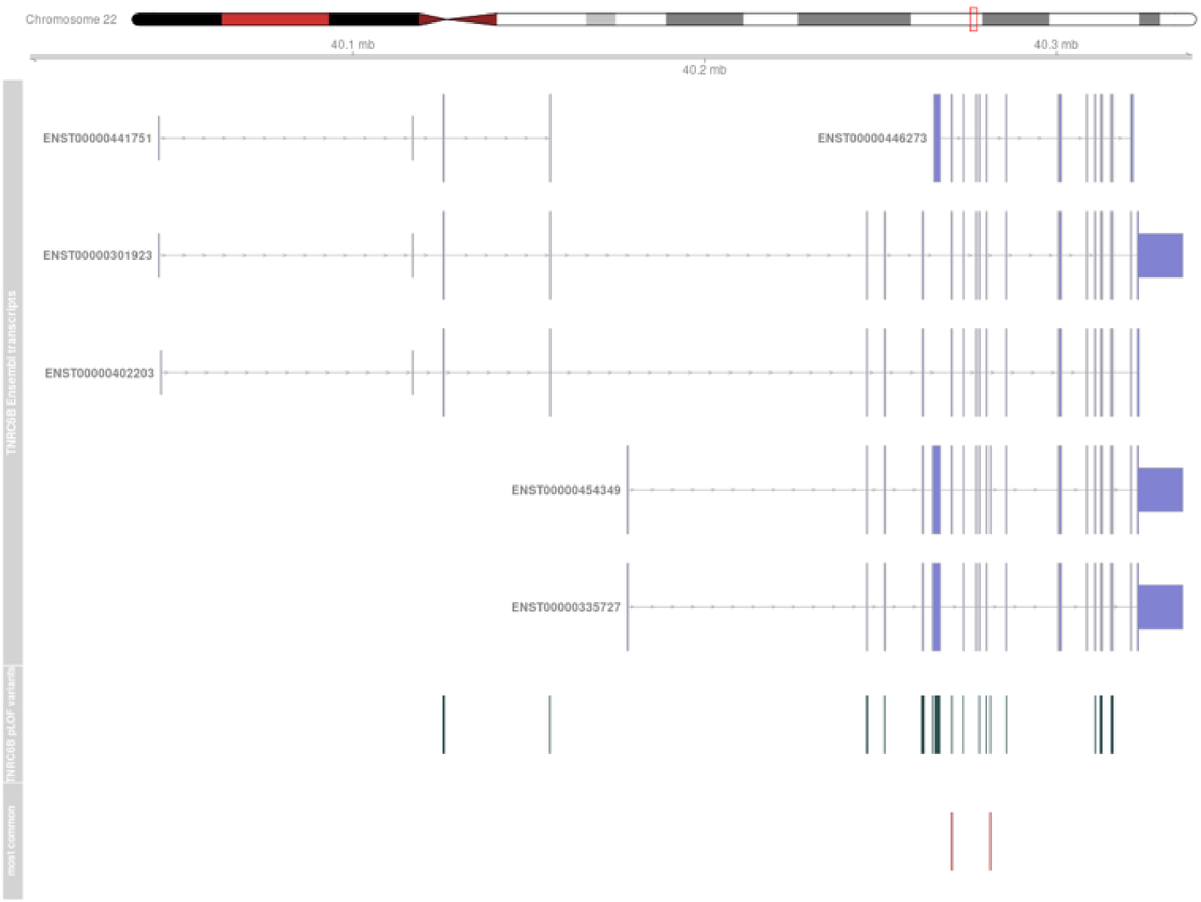
Chromosome 22
40.1 mb
40.2 mb
40.3 mb
TNRC6B Ensembl transcripts
ENST00000441751
ENST00000446273
ENST00000301923
ENST00000402203
ENST00000454349
ENST00000335727
TNRC6B pLOF variants
most common

#### Supplementary Figure 5: Association of drug target genes with HbA1c and T2D

Associations with HbA1c and T2D for variants in 8 genes encoding T2D drug targets (agonist targets: *GLP1R, IGF1R, PPARG, INSR*; antagonist targets: *SLC5A2, DPP4, KCNJ11, ABCC8*). The red line indicates the significance threshold ($p < 0.003$ correcting for 15 variant sets tested) and the blue line indicates nominal significance of $p < 0.05$.

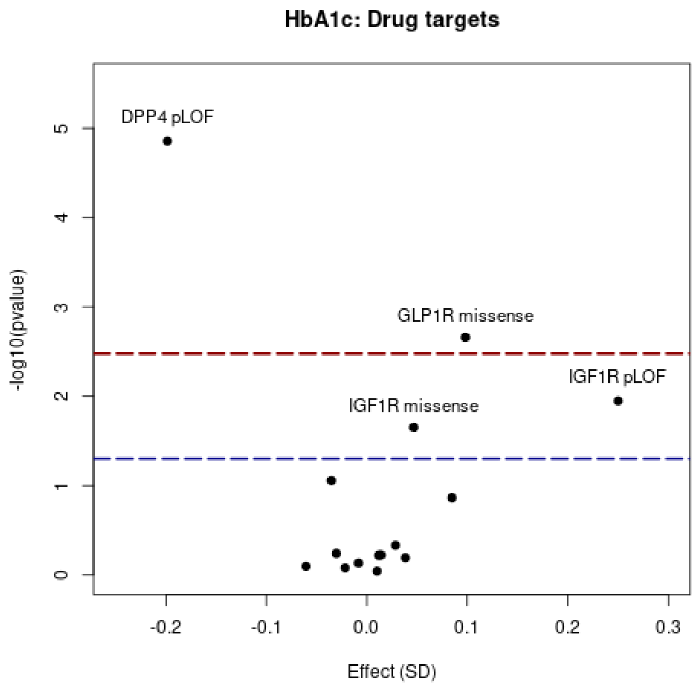


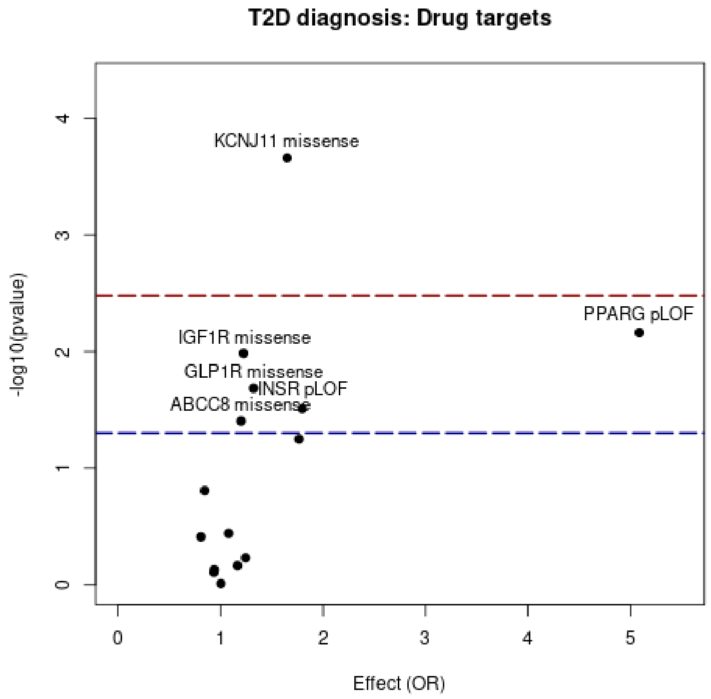

#### Supplementary Figure 6: Associations for genes near T2D GWAS loci

All genes in our study are colored grey and the two genes nearest T2D GWAS loci [1] are colored black. Red genes show significant associations with T2D or HbA1c in our study ($p < 2.41 \times 10^{-5}$). Genes shown twice have two variant sets (pLOF and missense) associating with the trait.

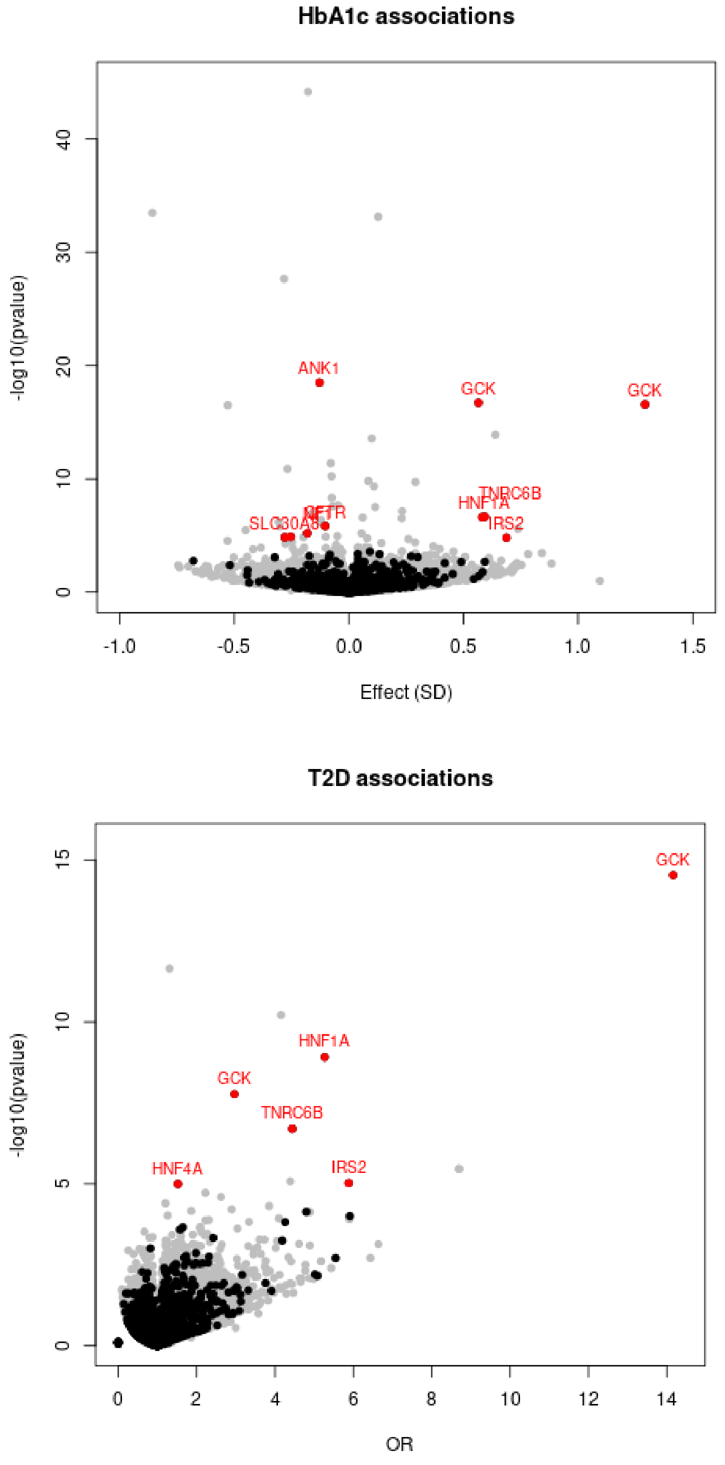

### Supplementary Tables

#### Supplementary Table 1: Numbers of pLOF and damaging missense variants in UKBB

Information for rare variants (MAF ≤ 1%) in each subpopulation of UKBB. CADD; combined annotation dependent depletion score.

| Population | N pLOF | N pLOF singletons | N genes with pLOF | N missense (CADD ≥ 25) | N missense (CADD ≥ 25) singletons | N genes with missense (CADD ≥ 25) | N total variant sets (pLOF+missense) |
|---|---|---|---|---|---|---|---|
| All | 726,422 | 425,081 | 16,477 | 2,139,471 | 1,060,539 | 17,312 | 33,789 |
| White | 574,950 | 338,916 | 16,348 | 1,691,784 | 838,024 | 17,222 | 33,570 |
| Black | 38,888 | 27,955 | 8,082 | 174,550 | 120,895 | 14,645 | 22,727 |
| Asian | 47,511 | 33,045 | 8,988 | 212,846 | 140,152 | 15,169 | 24,157 |
| Chinese | 9,336 | 6,841 | 2,987 | 53,486 | 39,008 | 9,909 | 12,896 |

#### Supplementary Table 2: T2D associations for participants recruited in England

| gene | Variant set | title | population | pvalue | OR | 95% CI - | 95% CI + | n cases | n carrier | n carrier cases | n expected |
|---|---|---|---|---|---|---|---|---|---|---|---|
| GCK | missense CADD ≥ 25 | T2D | English recruited | $1.38x10^{-8}$ | 3.11 | 2.1 | 4.6 | 22828 | 177 | 32 | 12.59 |
| GCK | pLOF | T2D | English recruited | $1.51x10^{-14}$ | 14.63 | 7.38 | 29 | 22828 | 37 | 18 | 2.63 |
| GIGYF1 | pLOF | T2D | English recruited | $1.24x10^{-10}$ | 4.33 | 2.77 | 6.76 | 22828 | 111 | 27 | 7.89 |
| HNF1A | pLOF | T2D | English recruited | $4.05x10^{-10}$ | 5.64 | 3.28 | 9.7 | 22828 | 67 | 20 | 4.77 |
| PAM | missense CADD ≥ 25 | T2D | English recruited | $6.34x10^{-13}$ | 1.33 | 1.23 | 1.44 | 22828 | 8242 | 751 | 586.21 |
| TNRC6B | pLOF | T2D | English recruited | $6.30x10^{-7}$ | 4.39 | 2.45 | 7.85 | 22828 | 63 | 16 | 4.48 |

#### Supplementary Table 3: Conditional analysis for *GIGYF1* and *GCK* variants

| gene | variant set | title | adjustment | pvalue | effect (beta/OR) | 95% CI - | 95% CI + |
|---|---|---|---|---|---|---|---|
| GIGYF1 | pLOF | glucose | GCK pLOF and missense | $4.26x10^{-12}$ | 0.62 | 0.44 | 0.79 |
| GIGYF1 | pLOF | HbA1c | GCK pLOF and missense | $1.2x10^{-14}$ | 0.64 | 0.48 | 0.80 |
| GIGYF1 | pLOF | T2D | GCK pLOF and missense | $5.83x10^{-11}$ | 4.16 | 2.72 | 6.38 |

#### Supplementary Table 4: Glucose, Hba1c and T2D associations for all significant variant sets

For all variant sets significantly associated with at least one diabetes-related trait in our primary analysis, associations with the other traits are shown.

| Gene | Variant set | Pvalue glucose | Beta glucose | Pvalue hba1c | Beta hba1c | Pvalue T2D | OR T2D |
|---|---|---|---|---|---|---|---|
| GCK | pLOF | 1.56E-09 | 0.999 | 2.64E-17 | 1.292 | 2.96E-15 | 14.16 |
| HNF1A | pLOF | 0.01 | 0.317 | 2.14E-07 | 0.592 | 1.23E-09 | 5.27 |
| GIGYF1 | pLOF | 4.42E-12 | 0.616 | 1.28E-14 | 0.639 | 6.14E-11 | 4.15 |
| GCK | missense CADD ≥ 25 | 6.15E-11 | 0.487 | 1.86E-17 | 0.565 | 1.7E-08 | 2.96 |
| PAM | missense CADD ≥ 25 | 0.92 | 0.001 | 0.009 | 0.026 | 2.26E-12 | 1.31 |
| IRS2 | pLOF | 0.07 | 0.320 | 1.58E-05 | 0.687 | 9.45E-06 | 5.88 |
| TNRC6B | pLOF | 4.01E-05 | 0.507 | 2.36E-07 | 0.582 | 2E-07 | 4.44 |
| PDX1 | missense CADD ≥ 25 | 0.02 | 0.029 | 2.54E-07 | 0.060 | 3.99E-05 | 1.21 |
| PFAS | missense CADD ≥ 25 | 0.32 | 0.009 | 2.09E-08 | -0.048 | 4.43E-04 | 0.88 |
| APOB | pLOF | 0.36 | 0.043 | 6.94E-08 | 0.232 | 0.02 | 1.42 |
| PLD1 | pLOF | 0.14 | 0.073 | 2.99E-07 | 0.231 | 0.01 | 1.50 |
| PTPRH | pLOF | 0.69 | 0.008 | 4.39E-10 | 0.109 | 0.10 | 1.12 |
| BLVRB | missense CADD ≥ 25 | 0.49 | 0.010 | 2.53E-08 | -0.071 | 0.30 | 0.95 |
| PLA2G12A | missense CADD ≥ 25 | 0.39 | 0.011 | 5.74E-11 | -0.076 | 0.09 | 0.92 |
| AMPD3 | missense CADD ≥ 25 | 0.82 | 0.003 | 7.28E-34 | 0.127 | 0.42 | 1.04 |
| CARHSP1 | missense CADD ≥ 25 | 0.86 | -0.005 | 6.78E-07 | -0.127 | 0.18 | 0.86 |
| LCAT | missense CADD ≥ 25 | 0.81 | -0.011 | 1.29E-11 | -0.268 | 0.31 | 0.83 |
| RHAG | pLOF | 0.10 | -0.126 | 3.31E-34 | -0.858 | 0.21 | 0.64 |
| ANK1 | missense CADD ≥ 25 | 0.46 | -0.012 | 3.12E-19 | -0.129 | 0.31 | 0.94 |
| HK1 | missense CADD ≥ 25 | 0.39 | 0.028 | 1.08E-07 | -0.159 | 0.14 | 1.19 |
| AXL | missense CADD ≥ 25 | 0.93 | 0.001 | 4.11E-12 | -0.080 | 0.28 | 0.95 |
| SLC4A1 | missense CADD ≥ 25 | 0.31 | 0.032 | 1.76E-07 | -0.154 | 0.35 | 1.12 |
| TFR2 | missense CADD ≥ 25 | 0.73 | 0.009 | 4.24E-07 | -0.124 | 0.45 | 0.92 |
| TMC8 | missense CADD ≥ 25 | 0.25 | -0.026 | 3.01E-08 | 0.114 | 0.41 | 0.93 |
| EPB42 | pLOF | 0.46 | -0.050 | 6.11E-07 | -0.307 | 0.42 | 1.21 |
| TNFRSF13B | missense CADD ≥ 25 | 0.98 | 0.000 | 4.63E-09 | -0.075 | 0.71 | 0.98 |
| PFKM | missense CADD ≥ 25 | 0.52 | 0.018 | 2.16E-28 | -0.283 | 0.27 | 0.88 |

| | | | | | | | |
|---|---|---|---|---|---|---|---|
| EPB41 | pLOF | 0.58 | 0.038 | 3.14E-17 | -0.529 | 0.26 | 1.30 |
| PFKL | missense CADD ≥ 25 | 0.82 | 0.003 | 2.69E-14 | 0.100 | 0.86 | 1.01 |
| APEH | missense CADD ≥ 25 | 0.30 | 0.052 | 1.86E-10 | 0.290 | 0.96 | 1.01 |
| G6PC2 | missense CADD ≥ 25 | 4.62E-83 | -0.266 | 6.71E-45 | -0.179 | 0.97 | 1.00 |
| PLD1 | missense CADD ≥ 25 | 0.54 | 0.009 | 1.51E-10 | 0.084 | 0.89 | 1.01 |
| PIEZO1 | missense CADD ≥ 25 | 0.05 | -0.013 | 9.9E-133 | -0.147 | 0.52 | 1.02 |

#### Supplementary Table 5: OMIM disease associations for genes with a glucose, HbA1c or T2D association

Genes associated with glucose, HbA1c and/or T2D that are implicated in Mendelian disease according to OMIM [2].

| gene | Variant set | OMIM ID | OMIM Phenotype |
|---|---|---|---|
| AMPD3 | missense CADD ≥ 25 | 102772 | [AMP deaminase deficiency, erythrocytic] |
| ANK1 | missense CADD ≥ 25 | 612641 | Spherocytosis, type 1 |
| APOB | pLOF | 107730 | Hypobetalipoproteinemia : Hypercholesterolemia, familial, 2 |
| EPB41 | pLOF | 130500 | Elliptocytosis-1 |
| EPB42 | pLOF | 177070 | Spherocytosis, type 5 |
| GCK | pLOF | 138079 | MODY, type II : Hyperinsulinemic hypoglycemia, familial, 3 : Diabetes mellitus, permanent neonatal : Diabetes mellitus, noninsulin-dependent, late onset |
| GCK | missense CADD ≥ 25 | 138079 | MODY, type II : Hyperinsulinemic hypoglycemia, familial, 3 : Diabetes mellitus, permanent neonatal : Diabetes mellitus, noninsulin-dependent, late onset |
| HK1 | missense CADD ≥ 25 | 142600 | Hemolytic anemia due to hexokinase deficiency : Retinitis pigmentosa 79 : Neuropathy, hereditary motor and sensory, Russe type |
| HNF1A | pLOF | 142410 | Renal cell carcinoma : MODY, type III : Hepatic adenoma, somatic : Diabetes mellitus, insulin-dependent, 20 |
| LCAT | missense CADD ≥ 25 | 606967 | Fish-eye disease : Norum disease |
| PDX1 | missense CADD ≥ 25 | 600733 | Lacticacidemia due to PDX1 deficiency : MODY, type IV : Pancreatic agenesis 1 |
| PFKL | missense CADD ≥ 25 | 171860 | Hemolytic anemia due to phosphofructokinase deficiency |
| PFKM | missense CADD ≥ 25 | 610681 | Glycogen storage disease VII |

| | | | |
|---|---|---|---|
| PIEZO1 | missense CADD ≥ 25 | 611184 | Lymphatic malformation 6 : Dehydrated hereditary stomatocytosis with or without pseudohyperkalemia and/or perinatal edema |
| PLD1 | missense CADD ≥ 25 | 602382 | Cardiac valvular defect, developmental |
| PLD1 | pLOF | 602382 | Cardiac valvular defect, developmental |
| RHAG | pLOF | 180297 | Overhydrated hereditary stomatocytosis : Anemia, hemolytic, Rh-null, regulator type |
| SLC4A1 | missense CADD ≥ 25 | 109270 | Spherocytosis, type 4 : [Blood group, Wright] : Renal tubular acidosis, distal, AR : [Blood group, Waldner] : Renal tubular acidosis, distal, AD : [Blood group, Swann] : Ovalocytosis, SA type : [Blood group, Froese] : Cryohydrocytosis : [Blood group, Diego] : [Malaria, resistance to] |
| TFR2 | missense CADD ≥ 25 | 604720 | Hemochromatosis, type 3 |
| TMC8 | missense CADD ≥ 25 | 605829 | Epidermodysplasia verruciformis 2 |
| TNFRSF13B | missense CADD ≥ 25 | 604907 | Immunoglobulin A deficiency 2 : Immunodeficiency, common variable, 2 |

#### Supplementary Table 6: HbA1c-associated genes and red blood cell traits (supplied as an excel file).

#### Supplementary Table 7: Glucose and HbA1c associations adjusted for BMI

| gene | Variant set | title | adjustment | pvalue | Effect (SD) | 95% CI - | 95% CI + | N carrier measured |
|---|---|---|---|---|---|---|---|---|
| G6PC2 | missense CADD ≥ 25 | glucose | unadjusted | 4.62E-83 | -0.27 | -0.29 | -0.24 | 5128 |
| G6PC2 | missense CADD ≥ 25 | glucose | BMI | 6.63E-84 | -0.26 | -0.29 | -0.24 | 5110 |
| GCK | missense CADD ≥ 25 | glucose | unadjusted | 6.15E-11 | 0.49 | 0.34 | 0.63 | 173 |
| GCK | missense CADD ≥ 25 | glucose | BMI | 3.32E-11 | 0.49 | 0.34 | 0.63 | 173 |
| GCK | pLOF | glucose | unadjusted | 1.56E-09 | 1 | 0.67 | 1.32 | 35 |
| GCK | pLOF | glucose | BMI | 3.82E-09 | 0.96 | 0.64 | 1.28 | 35 |
| GIGYF1 | pLOF | glucose | unadjusted | 4.42E-12 | 0.62 | 0.44 | 0.79 | 121 |
| GIGYF1 | pLOF | glucose | BMI | 1.02E-10 | 0.57 | 0.4 | 0.74 | 120 |

| | | | | | | | | |
|---|---|---|---|---|---|---|---|---|
| G6PC2 | missense CADD ≥ 25 | HbA1c | unadjusted | 6.71E-45 | -0.18 | -0.2 | -0.15 | 5574 |
| G6PC2 | missense CADD ≥ 25 | HbA1c | BMI | 3.42E-47 | -0.18 | -0.2 | -0.15 | 5555 |
| GCK | missense CADD ≥ 25 | HbA1c | unadjusted | 1.86E-17 | 0.56 | 0.43 | 0.69 | 201 |
| GCK | missense CADD ≥ 25 | HbA1c | BMI | 9.48E-18 | 0.55 | 0.43 | 0.68 | 201 |
| GCK | pLOF | HbA1c | unadjusted | 2.64E-17 | 1.29 | 0.99 | 1.59 | 38 |
| GCK | pLOF | HbA1c | BMI | 5.94E-17 | 1.24 | 0.95 | 1.53 | 38 |
| GIGYF1 | pLOF | HbA1c | unadjusted | 1.28E-14 | 0.64 | 0.48 | 0.8 | 129 |
| GIGYF1 | pLOF | HbA1c | BMI | 3.59E-13 | 0.59 | 0.43 | 0.75 | 128 |
| HNF1A | pLOF | HbA1c | unadjusted | 2.14E-07 | 0.59 | 0.37 | 0.82 | 68 |
| HNF1A | pLOF | HbA1c | BMI | 4.60E-08 | 0.61 | 0.39 | 0.82 | 68 |
| PDX1 | missense CADD ≥ 25 | HbA1c | unadjusted | 2.54E-07 | 0.06 | 0.04 | 0.08 | 6694 |
| PDX1 | missense CADD ≥ 25 | HbA1c | BMI | 2.88E-08 | 0.06 | 0.04 | 0.08 | 6673 |
| PFAS | missense CADD ≥ 25 | HbA1c | unadjusted | 2.09E-08 | -0.05 | -0.06 | -0.03 | 12621 |
| PFAS | missense CADD ≥ 25 | HbA1c | BMI | 5.11E-09 | -0.05 | -0.06 | -0.03 | 12573 |
| TNRC6B | pLOF | HbA1c | unadjusted | 2.36E-07 | 0.58 | 0.36 | 0.8 | 70 |
| TNRC6B | pLOF | HbA1c | BMI | 5.16E-07 | 0.55 | 0.33 | 0.76 | 70 |

#### Supplementary Table 8: T2D associations adjusted for BMI

| gene | variant set | title | adjustment | pvalue | OR | 95% CI - | 95% CI + | n cases |
|---|---|---|---|---|---|---|---|---|
| GCK | missense CADD ≥ 25 | T2D | unadjusted | 1.7E-08 | 2.96 | 2.03 | 4.32 | 24695 |
| GCK | missense CADD ≥ 25 | T2D | BMI | 2.07E-09 | 3.34 | 2.25 | 4.95 | 24504 |
| GCK | pLOF | T2D | unadjusted | 2.96E-15 | 14.16 | 7.33 | 27.34 | 24695 |
| GCK | pLOF | T2D | BMI | 4.17E-15 | 16.27 | 8.11 | 32.64 | 24504 |
| GIGYF1 | pLOF | T2D | unadjusted | 6.14E-11 | 4.15 | 2.71 | 6.37 | 24695 |
| GIGYF1 | pLOF | T2D | BMI | 1.38E-09 | 4.08 | 2.59 | 6.43 | 24504 |
| HNF1A | pLOF | T2D | unadjusted | 1.23E-09 | 5.27 | 3.08 | 9 | 24695 |
| HNF1A | pLOF | T2D | BMI | 2.94E-11 | 6.89 | 3.9 | 12.16 | 24504 |

| | | | | | | | | |
|---|---|---|---|---|---|---|---|---|
| PAM | missense CADD ≥ 25 | T2D | unadjusted | 2.26E-12 | 1.31 | 1.21 | 1.41 | 24695 |
| PAM | missense CADD ≥ 25 | T2D | BMI | 4.62E-14 | 1.35 | 1.25 | 1.46 | 24504 |
| TNRC6B | pLOF | T2D | unadjusted | 2.00E-07 | 4.44 | 2.53 | 7.79 | 24695 |
| TNRC6B | pLOF | T2D | BMI | 1.89E-07 | 4.84 | 2.67 | 8.75 | 24504 |

#### Supplementary Table 9: SAIGE-gene results for variant sets of interest

SAIGE-Gene was run in the White population including related individuals. Whether the association is significant at the thresholds used in our primary analysis ($p \leq 7.82 \times 10^{-7}$ for biomarkers and $p \leq 1.46 \times 10^{-6}$ for T2D) is indicated.

| **Gene** | **Variant set** | **title** | **pvalue** | **significant** |
|---|---|---|---|---|
| G6PC2 | missense CADD ≥ 25 | glucose | 1.30E-99 | Y |
| GCK | missense CADD ≥ 25 | glucose | 2.01E-11 | Y |
| GCK | pLOF | glucose | 6.56E-11 | Y |
| GIGYF1 | pLOF | glucose | 8.32E-13 | Y |
| G6PC2 | missense CADD ≥ 25 | HbA1c | 1.17E-62 | Y |
| GCK | missense CADD ≥ 25 | HbA1c | 3.20E-25 | Y |
| GCK | pLOF | HbA1c | 1.75E-20 | Y |
| GIGYF1 | pLOF | HbA1c | 1.92E-22 | Y |
| HNF1A | pLOF | HbA1c | 1.52E-08 | Y |
| PDX1 | missense CADD ≥ 25 | HbA1c | 9.05E-12 | Y |
| PFAS | missense CADD ≥ 25 | HbA1c | 2.00E-10 | Y |
| TNRC6B | pLOF | HbA1c | 6.85E-06 | N |
| GCK | missense CADD ≥ 25 | T2D | 2.33E-09 | Y |
| GCK | pLOF | T2D | 3.27E-15 | Y |
| GIGYF1 | pLOF | T2D | 7.34E-09 | Y |
| HNF1A | pLOF | T2D | 3.29E-09 | Y |
| PAM | missense CADD ≥ 25 | T2D | 5.60E-14 | Y |
| TNRC6B | pLOF | T2D | 4.77E-05 | N |

#### Supplementary Table 10: Glucose and HbA1c associations excluding diagnosed diabetics

Burden tests for glucose and HbA1c were performed for variants sets of interest excluding individuals diagnosed with any form of diabetes (ICD10 codes E10, E11, E12, E13 and E14).

| **gene** | **variant set** | **title** | **pvalue** | **Beta (SD)** | **95% CI -** | **95% CI +** | **n carrier measured** |
|---|---|---|---|---|---|---|---|
| G6PC2 | missense | glucose | 4.62E-83 | -0.27 | -0.29 | -0.24 | 5128 |
| G6PC2 | missense | glucose non-diabetic | 9.03E-97 | -0.28 | -0.31 | -0.25 | 4758 |
| GCK | missense | glucose | 6.15E-11 | 0.49 | 0.34 | 0.63 | 173 |
| GCK | missense | glucose non-diabetic | 2.42E-07 | 0.40 | 0.25 | 0.55 | 143 |
| GCK | pLOF | glucose | 1.56E-09 | 1.00 | 0.67 | 1.32 | 35 |
| GCK | pLOF | glucose non-diabetic | 0.02 | 0.47 | 0.07 | 0.87 | 20 |
| GIGYF1 | pLOF | glucose | 4.42E-12 | 0.62 | 0.44 | 0.79 | 121 |
| GIGYF1 | pLOF | glucose non-diabetic | 2.95E-08 | 0.53 | 0.34 | 0.72 | 92 |

| | | | | | | | |
|---|---|---|---|---|---|---|---|
| G6PC2 | missense | HbA1c | 6.71E-45 | -0.18 | -0.20 | -0.15 | 5574 |
| G6PC2 | missense | HbA1c non-diabetic | 1.45E-55 | -0.19 | -0.21 | -0.17 | 5171 |
| GCK | missense | HbA1c | 1.86E-17 | 0.56 | 0.43 | 0.69 | 201 |
| GCK | missense | HbA1c non-diabetic | 5.46E-11 | 0.44 | 0.31 | 0.57 | 166 |
| GCK | pLOF | HbA1c | 2.64E-17 | 1.29 | 0.99 | 1.59 | 38 |
| GCK | pLOF | HbA1c non-diabetic | 5.95E-05 | 0.79 | 0.41 | 1.18 | 19 |
| GIGYF1 | pLOF | HbA1c | 1.28E-14 | 0.64 | 0.48 | 0.80 | 129 |
| GIGYF1 | pLOF | HbA1c non-diabetic | 8.29E-07 | 0.43 | 0.26 | 0.60 | 99 |
| HNF1A | pLOF | HbA1c | 2.14E-07 | 0.59 | 0.37 | 0.82 | 68 |
| HNF1A | pLOF | HbA1c non-diabetic | 0.013 | 0.30 | 0.06 | 0.54 | 50 |
| PDX1 | missense | HbA1c | 2.54E-07 | 0.06 | 0.04 | 0.08 | 6694 |
| PDX1 | missense | HbA1c non-diabetic | 4.54E-04 | 0.04 | 0.02 | 0.06 | 6132 |
| PFAS | missense | HbA1c | 2.09E-08 | -0.05 | -0.06 | -0.03 | 12621 |
| PFAS | missense | HbA1c non-diabetic | 1.9E-07 | -0.04 | -0.06 | -0.03 | 11790 |
| TNRC6B | pLOF | HbA1c | 2.36E-07 | 0.58 | 0.36 | 0.80 | 70 |
| TNRC6B | pLOF | HbA1c non-diabetic | 0.009 | 0.31 | 0.08 | 0.55 | 52 |

#### Supplementary Table 11: Glucose and HbA1c associations from primary care data

Associations with glucose and HbA1c measurements obtained from primary care (GP) data. Whether the association is significant adjusting for the number of variant sets and traits tested ($p \leq 0.004$) is indicated.

| gene | Variant set | title | pvalue | beta | 95% CI - | 95% CI + | N carrier measured | significant |
|---|---|---|---|---|---|---|---|---|
| GIGYF1 | pLOF | Glucose GP | 2.1E-06 | 0.65 | 0.38 | 0.92 | 50 | Y |
| GCK | pLOF | Glucose GP | 1.41E-08 | 1.29 | 0.85 | 1.74 | 18 | Y |
| GCK | missense | Glucose GP | 0.00253 | 0.36 | 0.13 | 0.59 | 67 | Y |
| G6PC2 | missense | Glucose GP | 1.6E-20 | -0.19 | -0.23 | -0.15 | 2192 | Y |
| G6PC2 | missense | HbA1c GP | 4.14E-06 | -0.13 | -0.19 | -0.08 | 1169 | Y |
| GCK | pLOF | HbA1c GP | 3.9E-06 | 1.12 | 0.65 | 1.60 | 16 | Y |
| GCK | missense | HbA1c GP | 0.0625 | 0.26 | -0.01 | 0.53 | 50 | N |
| GIGYF1 | pLOF | HbA1c GP | 1.19E-05 | 0.74 | 0.41 | 1.07 | 33 | Y |
| HNF1A | pLOF | HbA1c GP | 5.68E-04 | 0.65 | 0.28 | 1.01 | 27 | Y |
| PDX1 | missense | HbA1c GP | 0.003 | 0.08 | 0.03 | 0.13 | 1385 | Y |
| PFAS | missense | HbA1c GP | 0.001 | -0.06 | -0.10 | -0.02 | 2679 | Y |
| TNRC6B | pLOF | HbA1c GP | 0.01 | 0.58 | 0.13 | 1.03 | 18 | N |

#### Supplementary Table 12: T2D associations for novel genes in Geisinger Health System

For novel genes associated with diabetes traits in our primary analysis we tested relevant variant sets for association with T2D diagnosis in Geisinger Health System (GHS). Variant set definitions and analysis methods varied somewhat from our primary analysis (see Methods).

| Phenotype | Gene | Variant set | pvalue | OR (95% CI) | MAF (%) | Genotype counts cases, RR\|RA\|AA genotypes | Genotype counts controls, RR\|RA\|AA genotypes |
|---|---|---|---|---|---|---|---|
| T2D | GIGYF1 | pLOF; MAF < 1% | 0.01 | 1.8 (1.1, 2.8) | 0.05% | 25,808\|38\|0 | 63,697\|52\|0 |
| T2D | TNRC6B | pLOF; MAF < 1% | 0.66 | 1.1 (0.8, 1.4) | 0.16% | 25,775\|71\|0 | 63,531\|218\|0 |
| T2D | PFAS | pLOF plus deleterious missense; MAF < 1% | 0.29 | 1 (0.96, 1.1) | 1.78% | 24,898\|946\|2 | 61,512\|2,233\|4 |

#### Supplementary Table 13: Association results when singletons are excluded

| gene | Variant set | Included variants | phenotype | pvalue | effect (beta/OR) | n carrier | significant |
|---|---|---|---|---|---|---|---|
| GCK | missense CADD ≥ 25 | All | T2D | 1.70E-08 | 2.96 | 202 | Y |
| GCK | missense CADD ≥ 25 | No singletons | T2D | 3.04E-04 | 2.29 | 166 | N |
| GCK | pLOF | All | T2D | 2.96E-15 | 14.16 | 40 | Y |
| GCK | pLOF | No singletons | T2D | 2.57E-07 | 10.88 | 21 | Y |
| GIGYF1 | pLOF | All | T2D | 6.14E-11 | 4.15 | 131 | Y |
| GIGYF1 | pLOF | No singletons | T2D | 2.91E-05 | 3.64 | 72 | N |
| HNF1A | pLOF | All | T2D | 1.23E-09 | 5.27 | 73 | Y |
| HNF1A | pLOF | No singletons | T2D | 2.34E-08 | 5.21 | 63 | Y |
| PAM | missense CADD ≥ 25 | All | T2D | 2.26E-12 | 1.31 | 9357 | Y |
| PAM | missense CADD ≥ 25 | No singletons | T2D | 4.1E-12 | 1.30 | 9257 | Y |
| TNRC6B | pLOF | All | T2D | 2E-07 | 4.44 | 71 | Y |
| TNRC6B | pLOF | No singletons | T2D | 0.69 | 1.36 | 23 | N |
| G6PC2 | missense CADD ≥ 25 | All | glucose | 4.62E-83 | -0.27 | 5128 | Y |
| G6PC2 | missense CADD ≥ 25 | No singletons | glucose | 1.12E-82 | -0.27 | 5103 | Y |
| GCK | missense CADD ≥ 25 | All | glucose | 6.15E-11 | 0.49 | 173 | Y |
| GCK | missense CADD ≥ 25 | No singletons | glucose | 6.98E-05 | 0.33 | 142 | N |
| GCK | pLOF | All | glucose | 1.56E-09 | 1.00 | 35 | Y |
| GCK | pLOF | No singletons | glucose | 0.0015 | 0.69 | 20 | N |
| GIGYF1 | pLOF | All | glucose | 4.42E-12 | 0.62 | 121 | Y |
| GIGYF1 | pLOF | No singletons | glucose | 1.54E-05 | 0.52 | 66 | N |
| G6PC2 | missense CADD ≥ 25 | All | HbA1c | 6.71E-45 | -0.18 | 5574 | Y |
| G6PC2 | missense CADD ≥ 25 | No singletons | HbA1c | 9.98E-45 | -0.18 | 5548 | Y |
| GCK | missense CADD ≥ 25 | All | HbA1c | 1.86E-17 | 0.56 | 201 | Y |
| GCK | missense CADD ≥ 25 | No singletons | HbA1c | 6.13E-09 | 0.43 | 165 | Y |
| GCK | pLOF | All | HbA1c | 2.64E-17 | 1.29 | 38 | Y |
| GCK | pLOF | No singletons | HbA1c | 1.16E-05 | 0.90 | 21 | N |
| GIGYF1 | pLOF | All | HbA1c | 1.28E-14 | 0.64 | 129 | Y |

| | | | | | | | |
|---|---|---|---|---|---|---|---|
| GIGYF1 | pLOF | No singletons | HbA1c | 5.47E-09 | 0.65 | 72 | Y |
| HNF1A | pLOF | All | HbA1c | 2.14E-07 | 0.59 | 68 | Y |
| HNF1A | pLOF | No singletons | HbA1c | 4.75E-06 | 0.56 | 59 | N |
| PDX1 | missense CADD ≥ 25 | All | HbA1c | 2.54E-07 | 0.06 | 6694 | Y |
| PDX1 | missense CADD ≥ 25 | No singletons | HbA1c | 4.75E-07 | 0.06 | 6662 | Y |
| PFAS | missense CADD ≥ 25 | All | HbA1c | 2.09E-08 | -0.05 | 12621 | Y |
| PFAS | missense CADD ≥ 25 | No singletons | HbA1c | 7.07E-08 | -0.05 | 12493 | Y |
| TNRC6B | pLOF | All | HbA1c | 2.36E-07 | 0.58 | 70 | Y |
| TNRC6B | pLOF | No singletons | HbA1c | 0.09 | 0.34 | 23 | N |

#### Supplementary Table 14: Associations for variants in *SLC30A8* and *MC4R*

We examined glucose, HbA1c and T2D associations for *SLC30A8* and *MC4R*, genes reported to associate with T2D by Flannick and colleagues [3].

| variant set | title | pvalue | effect (SD/OR) | 95% CI - | 95% CI + | n carrier measured | n carrier cases | n expected |
|---|---|---|---|---|---|---|---|---|
| SLC30A8 pLOF | HbA1c | 1.44E-05 | -0.28 | -0.41 | -0.15 | 213 | | |
| SLC30A8 pLOF | glucose | 0.003 | -0.20 | -0.33 | -0.07 | 209 | | |
| SLC30A8 pLOF | T2D | 0.03 | 0.42 | 0.20 | 0.91 | | 7 | 15.81 |
| SLC30A8 missense | HbA1c | 6.16E-06 | -0.18 | -0.26 | -0.10 | 554 | | |
| SLC30A8 missense | glucose | 2.51E-04 | -0.16 | -0.25 | -0.08 | 493 | | |
| SLC30A8 missense | T2D | 0.06 | 0.69 | 0.47 | 1.01 | 577 | 28 | 39.15 |
| SLC30A8 pLOF+missense | HbA1c | 8.64E-10 | -0.21 | -0.28 | -0.14 | 767 | | |
| SLC30A8 pLOF+missense | glucose | 2.71E-06 | -0.17 | -0.25 | -0.10 | 702 | | |
| SLC30A8 pLOF+missense | T2D | 0.005 | 0.61 | 0.44 | 0.86 | | 35 | 54.96 |
| MC4R missense | HbA1c | 0.19 | 0.04 | -0.02 | 0.10 | 872 | | |
| MC4R missense | glucose | 0.78 | 0.01 | -0.06 | 0.08 | 787 | | |
| MC4R missense | T2D | 0.27 | 1.15 | 0.90 | 1.46 | | 73 | 62.56 |

#### Supplementary Table 15: Drug target gene associations with HbA1c and T2D

HbA1c and T2D associations for pLOF and damaging missense variants in the genes encoding T2D drug targets.

**p < 0.003 (significance correcting for 15 variant sets tested)

*p<0.05 (nominal significance)

| Variant set | title | pvalue | Effect (beta/OR) | 95% CI - | 95% CI + | N carrier measured or N carrier cases/N expected | Significance |
|---|---|---|---|---|---|---|---|

| | | | | | | | |
|---|---|---|---|---|---|---|---|
| DPP4 pLOF | HbA1c | 1.4E-05 | -0.20 | -0.29 | -0.11 | 424 | ** |
| GLP1R missense | HbA1c | 0.002 | 0.10 | 0.04 | 0.16 | 869 | ** |
| IGF1R pLOF | HbA1c | 0.01 | 0.25 | 0.06 | 0.44 | 91 | * |
| IGF1R missense | HbA1c | 0.02 | 0.05 | 0.01 | 0.09 | 2145 | * |
| DPP4 missense | HbA1c | 0.09 | -0.04 | -0.08 | 0.01 | 2085 | |
| ABCC8 pLOF | HbA1c | 0.14 | 0.08 | -0.03 | 0.20 | 274 | |
| KCNJ11 missense | HbA1c | 0.47 | 0.03 | -0.05 | 0.10 | 584 | |
| SLC5A2 pLOF | HbA1c | 0.57 | -0.03 | -0.14 | 0.08 | 306 | |
| INSR missense | HbA1c | 0.60 | 0.01 | -0.04 | 0.07 | 1253 | |
| ABCC8 missense | HbA1c | 0.60 | 0.01 | -0.03 | 0.06 | 1718 | |
| INSR pLOF | HbA1c | 0.64 | 0.04 | -0.12 | 0.20 | 133 | |
| SLC5A2 missense | HbA1c | 0.73 | -0.01 | -0.06 | 0.04 | 1485 | |
| PPARG pLOF | HbA1c | 0.80 | -0.06 | -0.52 | 0.40 | 16 | |
| GLP1R pLOF | HbA1c | 0.83 | -0.02 | -0.22 | 0.18 | 86 | |
| PPARG missense | HbA1c | 0.90 | 0.01 | -0.16 | 0.18 | 119 | |
| KCNJ11 missense | T2D | 2.19E-04 | 1.65 | 1.26 | 2.15 | 63/42.27 | ** |
| PPARG pLOF | T2D | 0.007 | 5.08 | 1.56 | 16.54 | 4/1.09 | * |
| IGF1R missense | T2D | 0.01 | 1.22 | 1.05 | 1.43 | 181/152.18 | * |
| GLP1R missense | T2D | 0.02 | 1.32 | 1.04 | 1.67 | 78/61.94 | * |
| INSR pLOF | T2D | 0.03 | 1.80 | 1.06 | 3.06 | 16/9.16 | * |
| ABCC8 missense | T2D | 0.04 | 1.20 | 1.01 | 1.43 | 143/121.92 | * |
| PPARG missense | T2D | 0.06 | 1.77 | 0.98 | 3.17 | 13/8.41 | |
| INSR missense | T2D | 0.16 | 0.84 | 0.67 | 1.07 | 77/89.08 | |
| DPP4 missense | T2D | 0.36 | 1.08 | 0.92 | 1.27 | 159/148.32 | |
| SLC5A2 pLOF | T2D | 0.39 | 0.81 | 0.50 | 1.31 | 18/21.98 | |
| GLP1R pLOF | T2D | 0.58 | 1.24 | 0.57 | 2.72 | 7/5.97 | |
| IGF1R pLOF | T2D | 0.68 | 1.17 | 0.56 | 2.42 | 8/6.65 | |
| DPP4 pLOF | T2D | 0.74 | 0.94 | 0.65 | 1.37 | 30/30.53 | |
| ABCC8 pLOF | T2D | 0.78 | 0.94 | 0.58 | 1.50 | 19/19.54 | |
| SLC5A2 missense | T2D | 0.97 | 1.00 | 0.82 | 1.23 | 104/105.37 | |

#### Supplementary Table 16: *GIGYF1* pLOF associations with cholesterol levels

The association of *GIGYF1* pLOF with total and LDL cholesterol was tested 1) adjusting for use of cholesterol-lowering medication and 2) excluding those on cholesterol-lowering medication from the analysis.

| **gene** | **variant set** | **title** | **adjustment/subset** | **pvalue** | **beta** | **95% CI -** | **95% CI +** | **n carrier measured** |
|---|---|---|---|---|---|---|---|---|
| GIGYF1 | pLOF | total cholesterol | unadjusted | 2.44E-12 | -0.61 | -0.78 | -0.44 | 128 |
| GIGYF1 | pLOF | LDL cholesterol | unadjusted | 2.40E-10 | -0.56 | -0.73 | -0.38 | 128 |
| GIGYF1 | pLOF | total cholesterol | adjusted cholesterol medication | 4.14E-11 | -0.52 | -0.67 | -0.36 | 128 |

| | | | | | | | | |
|---|---|---|---|---|---|---|---|---|
| GIGYF1 | pLOF | LDL cholesterol | adjusted cholesterol medication | 4.68E-09 | -0.46 | -0.62 | -0.31 | 128 |
| GIGYF1 | pLOF | total cholesterol | exclude cholesterol medication | 3.70E-07 | -0.46 | -0.63 | -0.28 | 96 |
| GIGYF1 | pLOF | LDL cholesterol | exclude cholesterol medication | 1.54E-05 | -0.39 | -0.57 | -0.21 | 96 |

#### Supplementary Table 17: rs221783 is an eQTL for *GIGYF1*

GTEx eQTL data for rs221783 in selected tissues. The nominal p-value is shown along with the slope from the linear regression (giving direction of effect) and standard error (SE) for the T allele. In all cases, rs221783 is highly or completely correlated with the best eQTL for *GIGYF1* as shown in the "$R^2$ with best" column.

| **Tissue** | **rsid** | **GTEx variant id** | **gene name** | **pvalue** | **Slope (T allele)** | **Slope SE** | **rsid for best eQTL** | **$R^2$ with best** |
|---|---|---|---|---|---|---|---|---|
| Adipose - subcutaneous | rs221783 | chr7_100695291_T_C_b38 | *GIGYF1* | $1.57x10^{-30}$ | 0.35 | 0.03 | rs221781 | 1.00 |
| Lung | rs221783 | chr7_100695291_T_C_b38 | *GIGYF1* | $1.00x10^{-17}$ | 0.32 | 0.04 | rs221774 | 1.00 |
| Pancreas | rs221783 | chr7_100695291_T_C_b38 | *GIGYF1* | $6.40x10^{-10}$ | 0.38 | 0.06 | rs221786 | 0.96 |
| Thyroid | rs221783 | chr7_100695291_T_C_b38 | *GIGYF1* | $1.14x10^{-26}$ | 0.37 | 0.03 | rs221798 | 1.00 |

#### Supplementary Table 18: Conditional analysis of common variant and *GIGYF1* pLOF

Conditional analysis was performed in individuals from the White subset with array genotypes, exome sequencing data and biomarker measurements available.

| **phenotype** | **Variants(s)** | **adjustment** | **pvalue** | **Beta (SD)** |
|---|---|---|---|---|
| glucose | *GIGYF1* pLOF | adj rs221783 | 5.29E-12 | 0.71 |
| glucose | *GIGYF1* pLOF | unadjusted | 5.66E-12 | 0.71 |
| HbA1c | *GIGYF1* pLOF | adj rs221783 | 6.38E-12 | 0.65 |
| HbA1c | *GIGYF1* pLOF | unadjusted | 6.83E-12 | 0.65 |
| glucose | rs221783 | adj pLOF | 1.41E-10 | -0.03 |
| glucose | rs221783 | unadjusted | 1.51E-10 | -0.03 |
| HbA1c | rs221783 | adj pLOF | 9.26E-07 | -0.02 |
| HbA1c | rs221783 | unadjusted | 9.91E-07 | -0.02 |

#### Supplementary Table 19: Replication of rs221783 associations and meta-analysis

| **dataset** | **phenotype** | **chr** | **pos (hg19/hg38)** | **Ref (effect allele)** | **Alt** | **rsid** | **MAF** | **pvalue** | **effect in SD or OR (95% CI)** |
|---|---|---|---|---|---|---|---|---|---|
| UKBB | Glucose | 7 | 100292914/ 100695291 | T | C | rs221783 | 11.0% | $1.82x10^{-11}$ | -0.03 (-0.03, -0.02) |
| Biobank Japan | Glucose (blood sugar) | 7 | 100292914/ 100695291 | T | C | rs221783 | 3.5% | $1.71x10^{-4}$ | -0.05 (-0.07, -0.02) |

| | | | | | | | | | |
|---|---|---|---|---|---|---|---|---|---|
| UKBB+Japan | Glucose | 7 | 100292914/ 100695291 | T | C | rs221783 | - | $4.43 \times 10^{-14}$ | -0.03 (-0.04, -0.02) |
| UKBB | T2D (ICD10 E11) | 7 | 100292914/ 100695291 | T | C | rs221783 | 11.0% | 0.005 | 0.96 (0.93, 0.99) |
| FinnGen R3 | T2D (E4 DM2) | 7 | 100292914/ 100695291 | T | C | rs221783 | 15.4% | 0.02 | 0.96 (0.93, 0.99) |
| UKBB+FinnGen | T2D | 7 | 100292914/ 100695291 | T | C | rs221783 | - | $2.98 \times 10^{-4}$ | 0.99 (0.94, 0.98) |

#### Supplementary Table 20: Identification of causal genes at T2D GWAS loci

T2D and HbA1c associations for pLOF and missense variants in the two genes closest to T2D GWAS hits [1] were examined. Significant results are shown ($p \leq 2.41 \times 10^{-5}$ adjusting for 2071 variant sets tested).

| **Variant set** | **title** | **pvalue** | **effect (beta/OR)** | **95% CI -** | **95% CI +** |
|---|---|---|---|---|---|
| ANK1 missense | HbA1c | 3.12E-19 | -0.13 | -0.16 | -0.10 |
| GCK missense | HbA1c | 1.86E-17 | 0.56 | 0.43 | 0.69 |
| GCK pLOF | HbA1c | 2.64E-17 | 1.29 | 0.99 | 1.59 |
| HNF1A pLOF | HbA1c | 2.14E-07 | 0.59 | 0.37 | 0.82 |
| TNRC6B pLOF | HbA1c | 2.36E-07 | 0.58 | 0.36 | 0.80 |
| CFTR pLOF | HbA1c | 1.4E-06 | -0.10 | -0.15 | -0.06 |
| SLC30A8 missense | HbA1c | 6.16E-06 | -0.18 | -0.26 | -0.10 |
| NF1 pLOF | HbA1c | 1.27E-05 | -0.25 | -0.37 | -0.14 |
| SLC30A8 pLOF | HbA1c | 1.44E-05 | -0.28 | -0.41 | -0.15 |
| IRS2 pLOF | HbA1c | 1.58E-05 | 0.69 | 0.38 | 1.00 |
| GCK pLOF | T2D | 2.96E-15 | 14.16 | 7.33 | 27.34 |
| HNF1A pLOF | T2D | 1.23E-09 | 5.27 | 3.08 | 9.00 |
| GCK missense | T2D | 1.7E-08 | 2.96 | 2.03 | 4.32 |
| TNRC6B pLOF | T2D | 2.00E-07 | 4.44 | 2.53 | 7.79 |
| IRS2 pLOF | T2D | 9.45E-06 | 5.88 | 2.69 | 12.89 |
| HNF4A missense | T2D | 1.01E-05 | 1.52 | 1.26 | 1.84 |

### Supplementary Note

We examined the quality and location of individual variants in the following sets; *GIGYF1* pLOF, *TNRC6B* pLOF and *PFAS* damaging missense variants.

For *GIGYF1*, all pLOF variants overlapped both *GIGYF1* protein-coding transcripts. The most common *GIGYF1* pLOF variant in our data (rs770150936, hg38 7:100687545:CA:C, MAF = 0.0054%), carried by 39 people, is predicted to result in a frameshift and premature termination of translation at position 234 (ENSP00000275732.4:p.Leu111ArgfsTer234). We examined quality metrics for this variant and found that it was sequenced to a mean depth of 33x and had an allele quality score of 1142 (in 70th percentile for quality). The 7:100687545:CA:C variant is present in gnomAD v2.1.1 and v3 and has a frequency in the non-Finnish European population of 0.0048% and 0.0031% respectively.

For *PFAS*, the missense variants contributing most to the association with HbA1c are hg38 17:8258163:G:A (rs141777945, MAF = 0.2%) resulting in a glutamate to lysine substitution at position 434 (ENSP00000313490.6:p.Glu434Lys) and hg38 17:8262979:G:A (rs34939404, MAF = 0.7%) resulting in an alanine to threonine substitution at position 466 (ENSP00000313490.6:p.Ala466Thr). These variants were sequenced to a mean depth of 31x and 24x respectively and both had quality scores in the 75$^{th}$ percentile. Both are present at similar frequencies in gnomAD. These missense variants affect the longest protein-coding *PFAS* transcript ENST00000314666 (canonical and MANE select transcript).

Individual *TNRC6B* pLOF variants tended to be rare with the most common variants carried by just 9 (hg38 22:40270205:G:T; NP_001020014.1:p.Glu217Ter) and 7 (hg38 22:40281161:G:T NP_001155973.1:p.Glu1152Ter) individuals in White population respectively. This is consistent with the fact that the gene is constrained (pLI=1 and LOEUF=0.17). Both variants were sequenced to a mean depth of at least 42x and had high quality scores. Only 22:40281161:G:T is present in gnomAD and is flagged by the LOFTEE algorithm as potentially occurring in a misannotated exon (PHYLOCSF_UNLIKELY_ORF). Strikingly, 48 out of 71 carriers tested carried singleton variants. The large number of singletons together with the fact that *TNR6CB* is constrained suggest that some of these pLOF variants may not result in true loss of function or be due to sequencing errors.

#### Regeneron Genetics Center Banner Author List and Contribution Statements

All authors/contributors are listed in alphabetical order.

**RGC Management and Leadership Team**

Goncalo Abecasis, Aris Baras, Michael Cantor, Giovanni Coppola, Aris Economides, Luca A. Lotta, John D. Overton, Jeffrey G. Reid, Alan Shuldiner, Katia Karalis and Katherine Siminovitch, .

Contribution: All authors contributed to securing funding, study design and oversight. All authors reviewed the final version of the manuscript.

**Sequencing and Lab Operations**

Christina Beechert, Caitlin Forsythe, M.S., Erin D. Fuller, Zhenhua Gu, M.S., Michael Lattari, Alexander Lopez, M.S., John D. Overton, Thomas D. Schleicher, M.S., Maria Sotiropoulos Padilla, M.S., Louis Widom, Sarah E. Wolf, M.S., Manasi Pradhan, M.S., Kia Manoochehri, Ricardo H. Ulloa.

Contribution: C.B., C.F., A.L., and J.D.O. performed and are responsible for sample genotyping. C.B, C.F., E.D.F., M.L., M.S.P., L.W., S.E.W., A.L., and J.D.O. performed and are responsible for exome sequencing. T.D.S., Z.G., A.L., and J.D.O. conceived and are responsible for laboratory automation. M.S.P., K.M., R.U., and J.D.O are responsible for sample tracking and the library information management system.

**Genome Informatics**

Xiaodong Bai, Suganthi Balasubramanian, Andrew Blumenfeld, Boris Boutkov, Gisu Eom, Lukas Habegger, Alicia Hawes, B.S., Shareef Khalid, Olga Krasheninina, M.S., Rouel Lanche, Adam J. Mansfield,

B.A., Evan K. Maxwell, Mrunali Nafde, Sean O'Keeffe, M.S., Max Orelus, Razvan Panea, Tommy Polanco, B.A., Ayesha Rasool, M.S., Jeffrey G. Reid, William Salerno, Jeffrey C. Staples,

Contribution: X.B., A.H., O.K., A.M., S.O., R.P., T.P., A.R., W.S. and J.G.R. performed and are responsible for the compute logistics, analysis and infrastructure needed to produce exome and genotype data. G.E., M.O., M.N. and J.G.R. provided compute infrastructure development and operational support. S.B., S.K., and J.G.R. provide variant and gene annotations and their functional interpretation of variants. E.M., J.S., R.L., B.B., A.B., L.H., J.G.R. conceived and are responsible for creating, developing, and deploying analysis platforms and computational methods for analyzing genomic data.

**Clinical Informatics**

Michael Cantor and Dadong Li

Contribution: All authors contributed to developing the phenotype definitions.

**Translational Genetics:**

Adam Locke, Niek Verweij, Jonas Nielsen, Jonas Bovijn, Tanima De, Mary Haas, Parsa Akbari and Olukayode Sosina

Contribution: All authors contributed to the analytical analysis of the project.

**Research Program Management**

Marcus B. Jones, Jason Mighty, Michelle G. LeBlanc, and Lyndon J. Mitnaul,

Contribution: All authors contributed to the management and coordination of all research activities, planning and execution. All authors contributed to the review process for the final version of the manuscript.
